# Antiretroviral therapy adherence interventions among persons who use alcohol and other substances in fisherfolk communities: A systematic review

**DOI:** 10.1101/2024.08.16.24312118

**Authors:** Martin Osayande Agwogie, Landon Kuester, Polly Radcliffe, Andrea Gordon, Mary Loos, Anna Williams

## Abstract

**Background:** Suboptimal adherence to antiretroviral therapy (ART) is a major challenge in Human Immunodeficiency Virus (HIV) treatment interventions among persons who use alcohol and other substances (AOS) in fisherfolk communities. While studies have identified barriers to ART adherence and interventions to address these barriers in this population, no systematic review has been conducted to establish the significance of these interventions.

**Methods:** Searches were conducted in three electronic databases: PubMed, Web of Science and PsycINFO for studies conducted in English between 1996 and April 2024 to identify relevant primary studies that included adult males or females, engages in fishing activities, use alcohol or other substances and are HIV positive. Outcomes include any measure of adherence to ART, a reduction or abstinence from alcohol and other substance use and associated consequences that could hinder ART adherence.

**Results:** 54 studies were identified and screened against the inclusion/exclusion criteria. Five papers met the inclusion criteria (three quantitative designs, one qualitative design and one mixed methods design). Seven interventions were identified, these include counselling, peer support, screening and brief intervention, economic straightening, social network, gender transformative programmes and prescription monitoring.

**Conclusion:** Findings highlight the significance of alcohol use reduction interventions and gender transformative programmes particularly among men to encourage ART adherence in fisherfolk communities. To achieve the universal target of an end to the HIV scourge by 2030, specific hard to reach populations like fisherfolk communities with high HIV prevalence, alcohol and other substance use needs particular attention.

## 1. Introduction

Suboptimal adherence to antiretroviral therapy (ART) by people living with Human Immunodeficiency Virus (PLHIV) has remained a major barrier to effective HIV control particularly among persons who use alcohol and other substances (AOS) in fisherfolk communities (Kapesa et al., 2018; Sileo et al., 2019a; Tumwesigye et al., 2012). ART has been a global treatment for HIV with many lives saved (Parashar et al., 2016; Walter and Petry, 2016; Weber et al., 2013). However, positive outcomes depend on optimal ART adherence (O’Halloran Leach et al., 2021). Meeting optimal ART adherence has been a major challenge in HIV control (Fuge et al., 2022; Mohd Salleh et al., 2018; O’Halloran Leach et al., 2021). Moreover, achievements tend not to be uniform across nations, populations and settings (Parashar et al., 2016). For example, a systematic review reported that up to 75% of PLHIV on ART in the general population complied with treatment regimen (Detsis et al., 2017) with up to 92% viral load suppression (UNAIDS, 2022). However, in fishing communities, adherence is estimated below 70% (Sileo et al., 2019b) with less than 40% viral load suppression among persons who use AOS (Deryabina and El-Sadr, 2019). Similarly, while life expectancy among PLHIV is gradually levelling up with the general population, this is not the case among persons who use AOS (Marcus et al., 2016; McManus et al., 2012; Petoumenos and Law, 2016). In some countries, significant numbers of people, often the most marginalized (e.g., fishing communities) are being neglected (El-Bassel et al., 2014; Musumari et al., 2021; West et al., 2014). Therefore, the gains of ART have not fully extended to all populations, particularly persons who use AOS, in different work settings and communities including fisherfolk communities.

It appears to date no systematic review on interventions to address non-adherence to ART among fisherfolk communities with substance use issues has been conducted. Thus, the need for a systematic review exploring interventions to encourage ART adherence for HIV treatment among persons who use AOS in fisherfolk communities and to assess the effectiveness of the interventions.

### 1.1. Fisherfolk Communities

Fisherfolk communities are riverine or coastal communities that engage in fishing activities as an occupation and as means of livelihood. Fishing communities have been associated with high prevalence of AOS use and HIV risky behaviours (Choo et al., 2015; Fort et al., 2012; Tiberio et al., 2018). Alcohol consumption cuts across both male and females particularly among people living with HIV (Bonnevie et al., 2020; Kissling et al., 2005; Tumwesigye et al., 2012). Studies put HIV risks and prevalence among fishing communities at between three and 10 times greater than the general population (Kapesa et al., 2018; Michalopoulos et al., 2016; Tumwesigye et al., 2012; West et al., 2014).

### 1.2. Risk factors for AOS use and HIV infection in fisherfolk communities

Some risk factors linked with AOS use among the fishing population includes youth population, occupational culture and hazards, male dominant profession, family dysfunction, geographical area, stress, migratory and social isolation (Allison and Mvula, 2002; Fort et al., 2012; Kuteesa et al., 2020; Sileo et al., 2019a). Fishing has been described as a dangerous occupation which directly or indirectly impacts the health and well-being of fishermen (Percin et al., 2012). In the United States, commercial fishing is reported as one of the most hazardous occupations with a fatality rate estimated at over 40 times more than the national average in 2019 (Centre for Disease Control [CDC], 2023).

A qualitative study reported marginalization of fisherfolk communities in the larger society, common HIV risk behaviours such as unsafe drug injection practices, unprotected sex and acceptable drug use behaviours within the fishing communities as part of occupational social networks (West et al., 2014). Sustaining a thriving fishing business is associated with high risks including extra labour and stress. One study reported at certain times when profits are high and the lake is stormy, berthing sites experience higher levels of alcohol consumption, sex trading, substance use, and child labour (Wandera et al., 2021). Another study identified peer influence in addition to alcohol consumption as major factors associated with risky sexual behaviour among fisherfolk (Ford and Chamratrithirong, 2008).

### 1.3. Barriers to ART adherence

#### 1.3.1. AOS use

AOS use have been identified as barriers to engagement in HIV treatment and adherence to ART in fishing communities (El-Bassel et al., 2014; Sileo et al., 2019a; 2019b). A systematic review with meta-analysis reported that people who use alcohol were approximately 60% less likely to adhere to ART prescriptions compared to those who don’t use alcohol or use less (Hendershot et al., 2009). Similarly, UNAIDS reports that people who inject drugs (PWID) constitute 70% of HIV infections globally with new infections of up to 35 times more than adults who do not inject drugs (UNAIDS, 2022). Studies show close to half of those who use drugs amongst fisherfolk have a history of injecting drugs during fishing trips, longer stay at sea and engaged in unsafe injection practices in the past 30 days (Choo et al., 2015). The prevalence of HIV alone among PWID is estimated at between 18% and 25% (Deryabina and El-Sadr, 2019). The risk of HIV spread among PWID is directly through blood exchange from sharing needles and risky sexual behaviours under the influence of drugs (CDC, 2018; Plankey et al., 2007; Thiede et al., 2009). Active injecting drug use has been linked with non-adherent ART (Joseph et al., 2015). And fishing communities have been particularly associated with unsafe drug injection practices (Choo et al., 2015; West et al., 2014). Studies have reported persons who use AOS, particularly PWID are stigmatized, denigrated, marginalized and blamed for their HIV positive status, thereby constituting hindrance to ART adherence (Strathdee et al., 2012). Besides PWID, stigmatization is one of the socio–structural barriers to ART adherence in fisherfolk communities (Sileo et al., 2019b).

Despite the risk of HIV infection among persons who use AOS and as barriers to ART adherence in fishing communities, the prevention of AOS use among PLHIV has remained challenging (Kuteesa et al., 2022; Naigino et al., 2023; Petoumenos and Law, 2016; Womack and Justice, 2020). Increased intoxication has been reported from drug use and mortality risk compared to persons with negative HIV status (Justice et al., 2016). In particular, the control of HIV among PWID will remain challenging if the fundamental issues driving HIV transmission and promote lack of adherence to ART are not addressed in this community.

#### 1.3.2. Structural and social conditions

Recent studies identified structural peculiarities and social conditions including physical environment, occupational mobility, low quality of care and administrative protocol, poor linkage to care centres and lack of social support as contributing factors to non-optimal adherence to ART in fishing communities (Bogart et al., 2016; Ombere and Nyambedha, 2023; Rosen et al., 2020; Sileo, et al., 2019b). Better understanding of these structural and social factors and associated impacts will help to align policies and protocol for optimal utilization of ART in fishing communities. Other barriers include lack of integrated care that addresses substance use, HIV, other medical and co-occurring mental health conditions, psychosocial concerns and patients’ perspective of efficacy (Amin and Douaihy, 2018; Roberts, 2000).

### 1.4. Interventions to improve ART Adherence

The World Health Organisation (WHO) categorized the different adherence improvement interventions into five dimensions: health system/health care team related, social and economic related, patient related, therapy and condition related (WHO, 2003). Subsequently, studies have identified different interventions to encourage adherence to ART across these dimensions in different populations, settings, and subgroups.

**Table 1.** Interventions to improve ART Adherence.

|  | Interventions |
| --- | --- |
| i | Telephone-based support such as telephone messaging (daily or weekly) (Finitsis et al., 2014; Lester et al., 2010; Maitland et al., 2008; Mannheimer and Hirsch-Moverman, 2015; Robbins et al., 2013), telephone-delivered mindfulness training (Carey et al., 2020), telephone calls (Collier et al., 2005) and other active reminder devices (ARD) or technologies such as pagers and pillboxes with in-built timers and alarms (Chung et al., 2011; Hardy et al., 2011; Pop-Eleches et al., 2011; Sabin et al., 2010; Uzma et al., 2011). |
| ii | Cognitive behavioural therapy (CBT) such as individual, group and web-based (Duncan et al., 2012; Fisher et al., 2011; Holstad et al., 2011; Kalichman et al., 2011; Safren et al., 2012; Weber et al., 2004). |
| iii | Counselling (Chung et al., 2011; Kalichman et al., 2011; Mannheimer and Hirsch-Moverman, 2015; Pradier et al., 2003); modified or directly observed therapy (DOT) (Bärnighausen et al., 2012; Mannheimer and Hirsch-Moverman, 2015; Pearson et al., 2007; Sarna et al., 2008); motivational interviewing (DiIorio et al., 2008; Hill and Kavookjian, 2012; Naar-King et al., 2013). |
| iv | Education (Fisher et al., 2011; Kalichman et al., 2011; Parsons et al., 2007; Pradier et al., 2003; Rawlings et al., 2003). |
| v | A once-daily regimen compared to two or more daily dosage (Cooper et al., 2010; Parienti et al., 2007; Portsmouth et al., 2005). |
| vi | Behavioural economics, contingency management or incentives (Bien-Gund et al., 2021; Leeman et al., 2010; Rosen et al., 2007; Roy Paladhi et al., 2022; Sorensen et al., 2007). |
| vii | Treatment supporters including structured home-based ART delivery, visitation by a nurse or other health care providers (Berrien et al., 2004; Rich et al., 2012; Saberi et al., 2012; Thurman et al., 2010; Wang et al., 2010). |
| viii | Peer led mentoring and support (Purcell et al., 2007; Ruiz et al., 2010; Simoni et al., 2009). |
| ix | Nutritional support (Bärnighausen et al., 2012; Byron et al., 2008; Ndekha et al., 2009; Tirivayi and Groot, 2011). |
| x | Community social networks (Mendelsohn et al., 2012; Mannheimer and Hirsch-Moverman, 2015; Shushtari et al., 2023). |
| xi | Interventions focussing on mental health treatment (e.g., depression) (Pyne et al., 2011); Friendship Bench, a brief psychological intervention developed to bridge the mental health gap of persons living with HIV (Ouansafi et al., 2021); stigma reduction interventions (Singha et al., 2020) and harm reduction among persons who use AOS (Lappalainen et al., 2015). |

Some of these interventions for ART adherence have been evaluated among persons who use AOS. For example, in a systematic review, DOT was found to be effective in encouraging adherence to ART and recommended in the International Association of Physicians in AIDS Care (IAPAC) guideline (Ford et al., 2009; Mannheimer and Hirsch-Moverman, 2015). Once-a-day single tablet regimen or fewer pills was also found to promote optimal adherence among persons who use AOS (Mohd Salleh et al., 2018; Samet et al., 2005) and harm reduction among persons who use illicit opioids (Lappalainen et al., 2015).

Studies have also reported no significant positive outcomes in promoting adherence to ART with some interventions. For example, peer led, support and mentoring (Collier et al., 2005; Purcell et al., 2007; Simoni et al., 2007). Other interventions that have been found not to show significant improvement in ART adherence in different studies include education alone (Rawlings et al., 2003; Tuldrà et al., 2000), couples related interventions (Remien et al., 2005), interventions focused on treatment of depression alone (Pyne et al., 2011), CBT for adherence and depression (CBT-AD) (Simoni et al., 2013), telephone counselling (Abrahams et al., 2010), single tablet regimen (Dejesus et al., 2009; Parienti et., 2007) and mindfulness based stress management (Duncan et al,, 2012).

Studies addressing ART adherence seem to be inconsistent in findings depending on the target population and settings. However, studies report that combination and multifaceted interventions tailored towards individual’s needs and the at-risk populations seem most promising (Attonito et al., 2020; Mannheimer and Hirsch-Moverman, 2015; Mills et al., 2014). Socio-economic, environmental and cultural risk factors for substance use and HIV transmission in fishing communities are multiple and cross-cutting. Available studies have established high prevalence of AOS use and HIV in fishing communities, associated barriers to ART adherence and interventions to encourage ART adherence among those who use AOS. Despite these findings, there seems to be limited data on interventions to encourage adherence to ART in fishing communities and how barriers have been effectively addressed. Moreover, no systematic review has been conducted to establish these.

As ART expansion continues with recorded achievements in the control of the global HIV epidemic, barriers to adherence among persons who use AOS in fisherfolk communities need to be addressed so as not to undermine the progress made so far. This is particularly important in achieving the goal of an end to the HIV/AIDS scourge by 2030 (UNAIDS, 2014). Therefore, more studies are required to identify interventions that have been used to address these barriers to encourage adherence, whether they were effective in this population without assuming that ‘one-size-fits all’ or that similar interventions in other populations are sufficient. It is hoped that this review will help to identify different interventions to encourage ART adherence for HIV treatment among persons who use AOS in fisherfolk communities and to assess the effectiveness of the interventions. This will guide recommendations and suggestions for further studies.

## Methodology

### Research Design

This is a systematic review of primary studies on interventions to encourage ART among people living with HIV who use AOS in fisherfolk communities, identify the interventions and establish how effective are the interventions. The review is registered with PROSPERO, the prospective register for systematic review, (Registration: CRD42023429458). The outcomes of primary and secondary importance respectively are ART adherence interventions and interventions to encourage ART adherence in the population.

### Data Sources

An overview of related literature and topic as identified in journal articles contributed to the framing of the query. Searches were conducted across three electronic databases: PubMed, Web of Science and PsycINFO. The following search terms with Boolean operators were used: (“substance related disorders” OR “drug misuse”) AND (“fisheries” OR “fisherfolk communit*”

OR “fishing communit*” OR “fishing zone” OR “fishing village” OR “fish farm” OR “fishing area” OR “fisherfolk” OR “fishermen” OR “fisherwomen”) AND (“behavior therapy” OR “psychosocial intervention” OR “treatment” OR “therapeutics” OR “patient care” OR “encourage” OR “adherence” OR “promote” OR “treatment adherence and compliance” OR “medication adherence” OR “anti retroviral therapy” OR “hiv infections” OR “acquired immunodeficiency syndrome” OR “antiretroviral therapy, highly active” OR “prevent hiv transmission” OR “health education” OR “health promotion” OR “harm reduction”).

In addition, manual hand-searching of the reference list of included studies in the review was undertaken for relevant articles not identified through the database searches and key authors were contacted via email to determine whether there were relevant forthcoming papers that had not appeared in the searches. This is to ensure that no relevant articles were excluded.

### Inclusion/exclusion criteria

**Table 2:** Inclusion/exclusion criteria.

| **Inclusion** | **Exclusion** |
| --- | --- |
| **Participants**<br>Male or female, 18 years and above, engages in fishing activities, use AOS and are HIV positive.<br>**Interventions**<br>Interventions for inclusion were either of or a combination of pharmacotherapy, psychosocial, educational and support with the primary aim of promoting adherence to ART.<br>**Types of design** | Studies that do not include AOS use, people living with HIV and in fisherfolk communities.<br>Articles not published in English language or published before January 1996 when ART has not received global recognition for the treatment of HIV. Systematic reviews and meta-analysis. |
| Mixed methods, including quantitative and qualitative designs, randomized and non-randomized control trials were considered.<br>Publications that are primary studies.<br>**Year and language**<br>Only studies conducted between January 1996 and April 2024 and published in English language were included. 1996 was used considering the year ART received global recognition for the treatment of HIV.<br>**Outcomes**<br>Outcomes measure includes adherence to ART, a reduction or abstinence from AOS use and associated consequences that could hinder adherence to ART and reduction of HIV transmission (El-Bassel et al., 2014; Hlophe et al., 2023). | Studies that do not have specific outcomes to encourage ART adherence and studies that have participants below the age of 18 years where outcomes are not segregated by age. |

### Study selection

MOA independently extracted articles from the search outcomes after duplicates were removed. MOA, PR and LK independently screened the titles and abstracts against the inclusion and exclusion criteria to determine full texts to be reviewed for eligibility. Where there was eligibility disagreement, AG, ML and AW resolved the disagreements. MOA, PR and LK further assessed the included studies for eligibility by reading through the full text. Discrepancies were resolved by AG, ML and AW. The review process is shown in PRISMA flow diagram as Fig. 1.

**Fig. 1.**
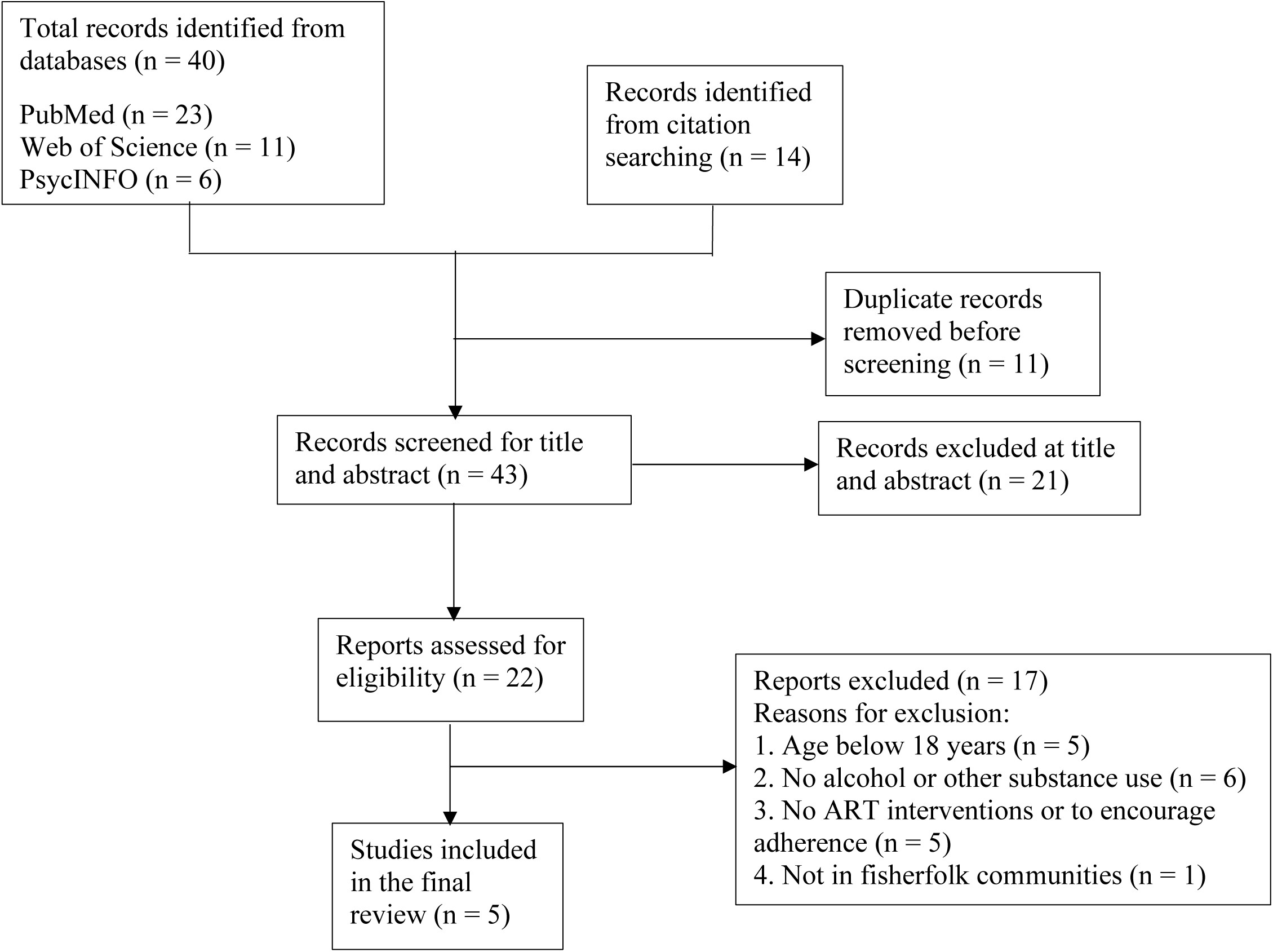
PRISMA flow diagram.

### Data extraction

The search outputs were imported into EndNote 2021 computer software for reference management and removal of duplicate references. A mixed methods data extraction form separated into qualitative and quantitative designs was used. MOA independently extracted descriptive and analytical data from the included studies (table 4). Extracted data includes author(s) and year of publication, title, country, age of respondents, gender, sample size, drugs involved, specific interventions, population target for intervention, type of design, outcome measure, index of adherence and study limitations. An agreement of data extraction was obtained from the authors. MOA then went through all the citations and extracted relevant data for accuracy. Where there were uncertainties, PR and LK were consulted. Once consensus was reached on the qualified studies, MOA entered relevant data into an excel data extraction form.

### Critical appraisal

The quality of studies was independently assessed by MOA and PR using methodological quality criteria outlined for qualitative, quantitative, and mixed method studies as provided in the Mixed Method Appraisal Tool Version 2018 (Hong et al., 2018). Each of these were assessed across five domains. For example, the qualitative studies were assessed by the following dimensions: a) if the qualitative approach were appropriate in answering the research question, b) adequacy of the qualitative data collection methods to address the research question, c) if findings were adequately derived from the data, d) if interpretation of results were sufficiently substantiated by the data and e) if there were coherence between data sources, collection, analysis, and interpretation. Similarly, quantitative studies were assessed on whether they were randomized control trials (RCT’s), non-randomized or descriptive studies. There were no randomized and non-randomized trials in the included studies, thus criteria for RCT’s were not utilized in this review. However, descriptive quantitative studies were assessed: a) if the sampling strategy were relevant to answering the research question, b) sample representativeness of the target population, c) appropriateness of measurements, d) level of risk of bias, and e) if statistical analysis were appropriate for the research question. Lastly, the mixed method studies were assessed for: a) rationale for using mixed method design, b) adequacy in the integration of the different components of the study to answer the research question, c) interpretation of outputs of all integrated components, d) resolution of divergence and inconsistences between the components and e) adherence to the quality criteria of each of the components. The final scores for the assessment were agreed through discussions between the primary investigator and the reviewers.

### Analysis

Qualitative analysis was undertaken by grouping key findings into main themes according to ART adherence intervention measures and encouraging adherence as relevant to the study by addressing barriers. First was the coding and categorization of first and second order empirical constructs while paying particular attention to the strengths and challenges of remote data collection practices and implications. This was followed by coding and categorization of study design (methodology, theoretical application, and synthesis), outcomes/findings and study limitations. All codes were presented in a MS excel table with their pertaining data and sorted into similarity to their potential themes. The themes used in this study were health system/health care team related, social and economic related, therapy related, patient related, condition related (WHO, 2003). Pooled prevalence and heterogeneity were not calculated due to the limited quantitative designs and similarity.

## Results

A total of 54 studies were found after running through the search strategy in the three databases (PubMed = 23, Web of Science = 11, PsycINFO = 6), citation searching (14) and follow up with authors (0). Eleven duplicate articles were removed. The process followed the Preferred Reporting Items for Systematic Reviews and Meta-Analysis (PRISMA) guidelines as reported in Fig. 1.

Out of the remaining 43 articles only 22 full-text studies met the criteria for screening and five papers from two studies were retained for inclusion in the analysis: quantitative studies (n = 3) (Kiene et al., 2019a; Sileo et al., 2019b; Sileo et al., 2021), qualitative studies (n = 1) (Sileo et al., 2019a), and mixed study (n = 1) (Sileo et al., 2019c). Other studies were excluded because of age (n = 5) (Bonnevie et al., 2020; Burgos-Soto et al., 2020; Kuteesa et al., 2022; Lubega et al., 2015; Mgabo et al., 2013), no alcohol or other substance use (n = 6)(Burke et al., 2017; Chang et al., 2016; Kagaayi et al., 2019; Long et al., 2017; Rosen et al., 2020; Sileo et al., 2019d), no ART interventions or to encourage adherence (n = 5)(Brown et al., 2017; Kiene et al., 2019b; Kissling et al., 2005; Ousley et al., 2018; Brown and George, 2019), and studies not in fisherfolk communities (n = 1)(Hickey et al., 2015) (see appendix 3).

Four out of the five papers in this review were from the same study (Sileo et al., 2019a; Sileo et al., 2019b; Sileo et al., 2019c; Sileo et al., 2021). In addition, the second study was conducted in the same country, Uganda (Kiene et al., 2019a). Though sample size for each of the four papers from one of the studies vary, the sample for the study was 300 males while the second study sampled 132 males and 168 females. Of the five papers, two (Sileo et al., 2019b; Kiene et al., 2019a) used structured questionnaires to collect data, one used structured interview, another one used in-depth semi-structured interview and one used both structured questionnaires and in-depth interview.

### Quality assessment

Results of the quality assessment are shown in Table 3. For each of the domains, the scores were reported in either “yes” or “no” and a percentage of number of “yeses” was used to report the final score in line with the Mixed method appraisal tool (Hong et al., 2018). All papers presented clear research questions, collected relevant data to address the questions, interpreted the results sufficiently from the data and adequately reported findings from the data. Rationales were provided for the use of both samples and designs. No ethical issues were identified in the papers. In summary, all the five papers had a score of 100% each demonstrating the high quality of the included papers.

**Table 3:** Quality assessment.

| **ID** | **Type of design** | **Mixed Methods Appraisal Tool** |
| --- | --- | --- |
| Sileo et al., 2019a | Qualitative design | 100 |
| Sileo et al., 2019b | Cross-sectional design | 100 |
| Sileo et al., 2019c | Mixed method (qualitative and cross-sectional) | 100 |
| Kiene et al., 2019a | Cross-sectional design | 100 |
| Sileo et al., 2021 | Cross-sectional design | 100 |

### Interventions

A total of seven interventions were identified from the review and are presented in six themes.

#### Counselling

Two papers identified counselling (Sileo et al., 2019a; Sileo et al., 2019b). Findings show that counselling on ART adherence and alcohol use helps to reduce the level of alcohol consumption with an increase in the proportion of prescription doses taken in the prior four days (Sileo et al., 2019a; Sileo et al., 2019b). The counselling services include providing information about AOS use and consequences, HIV transmission and consequences, knowledge about ART and benefits, sexuality education, allaying HIV associated fears and anxiety, discarding myths and misconceptions (Sileo et al., 2019a; Sileo et al., 2019b).

While earlier studies have identified counselling as effective intervention in promoting ART adherence (Kalichman et al., 2011; Mbuagbaw et al., 2015; Mills et al., 2014; Pradier et al., 2003), there have been some inconsistencies in findings (Hlophe et al., 2023; Reif et al., 2020).

The inconsistent findings may not be unconnected with the counselling strategies whether one on one, group or telephone, alone or in combination with other interventions which have been reported as important in making a difference. For example, Abrahams and colleagues noted that among the different counselling strategies, telephone counselling alone did not demonstrate significant positive outcomes in improving ART adherence (Abrahams et al., 2010) suggesting the need for a combination of interventions (Attonito et al., 2020; Brown and Adeagbo, 2022; Hlophe et al., 2023).

#### Gender transformative programmes

One of the papers highlighted the impacts of gender transformative strategies in improving ART adherence through improved patient retention (Sileo et al 2021). Gender transformative programmes work to challenge detrimental male gender norms and advance gender equality as a strategy to improve men and women’s health behaviours and outcomes (Sileo et al., 2021). Sileo and colleagues reported that gender transformative programmes restored masculine role of being seen as hard working, a provider to the family, husband, and sexual partner, possessing physical strength which were initially seen as being compromised by HIV positive diagnosis (Sileo et al., 2021). Studies have reported the influence of masculine norms and social expectations of gender roles in health behaviours (Baker et al., 2014; Fleming and Dworkin, 2016; WHO, 2007) and in HIV infection and management (Gottert, 2014; Shannon et al., 2012; Sileo, et al., 2019e) particularly in Sub-Sahara Africa (SSA). Reports in SSA have consistently maintained that men have worse ART treatment adherence compared to women and with greater mortality (Beckham et al., 2016; Cornell et al., 2011; Shand et al., 2014; Sileo et al., 2019e). For example, HIV diagnosis is viewed as a threat to men’s ego, social valued relationships, and their desires to be seen as strong, self-reliant, sexually successful and worthy of respect (Chikovore et al., 2016; Naugle et al., 2019; Okoror et al., 2016). Previous studies classified these roles under ‘reputation-based’ and ‘respectability-based’ masculine ideologies (Barker and Ricardo, 2005; Sikweyiya et al., 2014; Siu et al., 2013; Siu et al., 2014). Reputation-based gender roles relate to strength and sexual competence of men while respectability-based gender roles relate to being fatherly and provider for the family (Sileo et al., 2019c). Therefore, HIV stigma due to these norms may have significant effects on HIV care engagement and ART adherence for men who are adherent of the traditional masculine norms and are more likely to experience adverse impacts of stigma on their male self-worth, reputation and performance of their masculine ideals. Men with high level of internalized stigma have been reported to have increased likelihood of poor treatment adherence (Sileo et al., 2021). The fear of losing these identities and roles as well as the psychological impacts of being identified as HIV positive and seeking treatment, support the utilization and significance gender transformative programme. Earlier studies have reported that men who experienced ART healing effects on their health, strength, masculine identities, and role accomplishment adapted better to their HIV status and care engagement (Sileo et al., 2019c; Siu et al., 2014; Siu et al., 2012; Tsondai et al., 2017). However, for those who may experience some side effects such as dizziness and fatigue, the gender role of strength, hard work and provider becomes a barrier to ART adherence (Hlophe et al., 2023; Sileo et al., 2019c).

This finding has implications for the use of gender transformative interventions that includes stigma reduction strategies to address gender-based barriers to HIV treatment interventions among men. Previous studies have suggested the use of multi-level strategies that include individual level, structural and stigma reduction interventions. For example, Figueroa and colleagues used community dialogues to change community, gender and sexual norms in HIV prevention in Mozambique while expanding social support against HIV stigma (Figueroa et al., 2016) and have proved to be effective in reducing internalized stigma associated with HIV diagnosis and encourage adherence to ART (Pantelic et al., 2019; Sharma et al., 2017; Singh et al., 2020). Other gender transformative and structural-level interventions among PLHIV includes targeting HIV economic risk factors, such as cash transfer or microfinance programmes to reduce vulnerabilities to HIV infections and improve economic independence (Kilburn et al., 2018). A good understanding of the interacting and independent effects of endorsing inequitable gender norms and HIV stigma on HIV treatment interventions in this population could inform the advancement of gender specific HIV services to engage and retain fisherfolk and men in other key and hard to reach populations in HIV care.

#### Economic strengthening

One study reported the impact of commitment savings, as a component of economic strengthening, in reducing alcohol consumption and sexual HIV risky behaviours (Kiene et al., 2019a). Commitment savings was associated with lower odds of problematic alcohol use (Kiene et al., 2019a). Three reasons why commitment savings was effective in reducing sexual risk behaviour and alcohol use were identified. These includes, i) deterrent to impulsive purchase of alcohol and patronizing sex workers due to limited cash, ii) the small cost to accessing money which increases salience and iii) value of longer-term goals associated with savings and the overall benefits of achieving longer term life objectives (Kiene et al., 2019a).

The use of economic strengthening through commitment savings is an emerging intervention to encourage HIV care. Commitment savings involves promoting money saving, for example in a bank, cooperative or via mobile money with “soft commitment” by restricting access to money or charges fees for withdrawals (Kiene, et al., 2019a). Generally, economic strengthening interventions are aimed at improving the economic wellbeing of people through one or more of the following: financial literacy trainings, income generation, conditional cash transfers, micro savings and lending (Kiene et al., 2019a) and have been found to demonstrate positive effects on health outcomes (Ssewamala et al., 2012), reducing sexual risk behaviours (Witte et al., 2015) and improving ART adherence, treatment retention and quality of life measure in SSA (Bateganya et al., 2015). Commitment savings help to reduce impulse buying for people with issues of self-control and unnecessary cash withdrawals (Dupas and Robinson, 2013; Ky et al., 2016).

Prior studies have reported that people who use alcohol, particularly before sex are more likely to engage in an unprotected sex and get infected with HIV when compared with those who do not use alcohol or use at a lesser rate (Kiene et al., 2008; Woolf-King et al., 2011). And fishing communities operate on a cash-based economy where fisherfolk get paid daily in cash with limited mechanisms for savings (Sileo et al., 2016) thereby encouraging risky sexual behaviour and alcohol consumption (Keyes et al., 2011; Kuteesa et al., 2022; MacPherson et al., 2012).

While different forms of economic based strengthening interventions such as contingency management (Mbuagbaw et al., 2015) have been used to prevent high risk behaviours and encourage ART adherence, this use of commitment savings led economic strengthening seems to be novel. This seems to support the report of Mbuagbaw and colleagues who noted that ‘the role of theoretical underpinnings in adherence research for HIV is unclear’ (Mbuagbaw et al., 2015, 262). Therefore, adherence behaviour may go beyond the scope of any single theory. Innovative approaches in hard-to-reach populations are suggested. Moreover, complex interventions have not been shown to be more effective than simple ones (Chaiyachati et al., 2014).

Strong desires to save money for children and the future have been identified among PLHIV in previous studies (Sileo et al., 2016). However, a recent systematic review conducted in low and middle-income countries found that economic support was not statistically associated with ART adherence (Reif et al., 2020). Overall, this finding suggests that promoting money saving mechanisms which involves a commitment among fisherfolk may be particularly helpful for those who have problem with alcohol use in controlling the level of alcohol consumption and engagement in risky sexual behaviours. A randomized trial would be appropriate to determine if commitment saving behaviour in this population produces a reduction in AOS use that may encourage ART adherence.

#### Social network and peer support

A paper reported that social network and peer support increased ART adherence (Sileo et al., 2019c). Men were found to rely greatly on a network of male peers for instrumental and emotional support utilizing the unique working environment where men gather for work in a community setting.

One of the significant findings in this review is that men were found to rely greatly on a network of male peers for instrumental and emotional support thus utilizing the unique working environment where men gather for work in a community setting. People who engage in fishing activities are highly mobile, spend greater amount of time away from homes, mostly with peers and co-workers, and form community social networks. Previous studies have found community social networks (Enane et al., 2021; Mendelsohn et al., 2012; Mannheimer and Hirsch-Moverman, 2015), treatment and social supporters (Mills et al., 2014; Shushtari et al., 2023; van Wyk, 2021) and male peer-led groups (Baron et al., 2020; Bogart et al., 2020; Carbone et al., 2019, Holstad et al., 2012) as effective in encouraging ART adherence. Knowing other men on ART normalise HIV and reduce the fear of HIV associated stigma and discrimination, gives hope of longer life like colleague who may have lived with HIV for many years and still productive (Hlophe et al., 2023; Tsondai et al., 2017).

#### Prescription monitoring

In a cross-sectional study, Sileo and colleagues found that prescription monitoring enhances ART adherence with less proportion of missed ART in the prior four days (Sileo et al., 2019b). The Adult AIDS Clinical Trial Group (AACTG) scale was used to measure and monitor self-reported prescription adherence (Chesney et al., 2000). The use of directly observed therapy and monitoring have been recommended for persons who use AOS in previous studies (Mbuagbaw et al., 2015; Ford et al., 2009; Mannheimer and Hirsch-Moverman, 2015). While prescription monitoring enhances ART adherence it did not address alcohol use barrier to ART adherence (Sileo et al., 2019b).

#### Screening and brief intervention

Screening and brief intervention (SBI) helps to reduce alcohol consumption to enhance ART adherence and improve ART adherence (Sileo et al., 2019b). Respondents were found to be paying more attention to health needs, respect for health care providers and ART adherence. The successful implementation of SBI in reducing alcohol use is consistent with previous studies (Kaner et al., 2007). SBI involves universal screening for alcohol use, providing information, increasing motivation and teaching of behavioural change skills to reduce the level of alcohol use (Babor et al., 2007). Reductions in the level of alcohol consumption, paying attention to health needs and respect for health care providers suggests that SBI by care providers is acceptable to patients who use alcohol and are on ART (Sileo et al., 2019b). The inclusion of how these can help men to achieve their goals and fulfil masculine responsibilities including providing for families may increase the motivation for alcohol use reduction and cessation. However, Sileo and colleagues reported that there was no significant difference between those who had no record of drinking in past four days and those who engaged in low-risk drinking (Sileo et al., 2019b).

Thus, suggesting that higher consumption of alcohol may be driving suboptimal adherence.

Moreover, earlier studies demonstrated limited evidence in support of motivational interview based brief interventions to address alcohol use and HIV treatment engagement (Grimsrud et al., 2020; Sharma et al., 2017; Sileo et al., 2020). Further research is needed to test the effectiveness of SBI for fisherfolk who use AOS in a quantitative and randomized control trail. This will help to understand the needed best structure, delivery and environmental intervention in this unique population context.

The effects of the different interventions are summarized in table 4.

**Table 4:** Characteristics of included studies.

| ID | Title | Country | Age | Gender | Sample size | Type of Drugs | Intervention(s) | Target Population | Study Design | Outcome measure | Index of adherence | Limitations |
| --- | --- | --- | --- | --- | --- | --- | --- | --- | --- | --- | --- | --- |
| Sileo et al., 2019a | A qualitative study on alcohol consumption and HIV treatment adherence among men living with HIV in Ugandan fishing communities | Uganda | 18+ | Male | 30 | Alcohol | Counselling: This includes providing information on consequences of AOS use, reducing alcohol consumption, HIV transmission and consequences, knowledge about ART and benefits | Men in HIV care and on ART in a fishing communities | Qualitative design | Mean audit score 10.4<br>40% participants optimally adherent; 60% non optimally adherent i.e. taking less than 95% of doses of ART in four day recall period | Increase in the proportion of prescription doses taken in the prior four days | It is a qualitative design and purposive sampling method, therefore not generalizable to the larger population. Social desirability bias may have influenced the responses thus resulting in underreporting of the level of alcohol consumption and non-adherence to ART. |
| Sileo et al., 2019b | Substance use and its effect on antiretroviral treatment adherence among male fisherfolk living with HIV/AIDS in Uganda | Uganda | 18+ | Male | 300 | Alcohol and other substance use | Counselling: ART adherence counselling, allaying HIV associated fears and anxiety, discarding myths and misconception; Prescription monitoring with the use of the Adult AIDS Clinical Trial Group (AACTG) | Residents of fishing communities | Cross-sectional design | Self reported adherence to ART measure using AIDS Clinical Trial Group scale for hazardous drinking (AUDIT) and Index of current drinking (frequency and quantity) | The proportion of missed ART in the prior four days | As a cross-sectional design, there is no causal relationship between alcohol use and ART adherence. The accuracy of assessment of responses on alcohol use from the structured interview may have varied. Being a non-random sampling procedure, the outcome from the study cannot be generalized. By recruiting respondents directly from the clinic, the sample may have over- |
|  |  |  |  |  |  |  | scale to measure and monitor self-reported prescription adherence; Screening and brief intervention (SBI) which involves screening for alcohol use, increasing motivation and teaching of behavioural change skills to reduce the level of alcohol use. |  |  |  |  | represented the level of ART adherence. |
| Sileo et al., 2019c | Masculinity and engagement in HIV care among male fisherfolk on HIV treatment in Uganda | Uganda | 18+ | Male | 30 | Alcohol | Social network and peer-based interventions: Using a network of male peers for instrumental and emotional support. | Fisherfolk on ART | Mixed method (qualitative and cross-sectional) | Self reported adherence to ART measure using AIDS Clinical Trial Group scale for hazardous drinking (AUDIT) and Index of current drinking (frequency and quantity) Adherence ART<br>reported proportion of ARVs taken in the prior four days, out of the total ARVs prescribed.<br>Masculinity - narrative reports of 3 roles that men felt were the most important to fulfil now or the future<br>Self reported relation between HIV diagnosis and sense of manhood<br>Self reported relation between HIV diagnosis and ability to fulfil each of the roles | Increase participation in HIV care and adherence to ART | The generalizability of findings is limited, given the qualitative and cross-sectional nature of the study. The use of purposive sampling method also limits generalizability of the study. |
| Kiene et al., 2019a | Exploring the potential of savings-led economic strengthening HIV | Uganda | 18+ | Male and Female | 300 | Alcohol | Economic strengthening: Commitment savings that involves promoting | Residents of fishing communities | Cross-sectional design | Number of risky unprotected sex acts measured as the number | Commitment savings was associated with lower | Only 20% of the participants reported problematic use of alcohol. Commitment savings variable was not |
|  | interventions among high-risk economically vulnerable fishing communities in Uganda: Associations between use of commitment savings, sexual risk behavior, and problematic alcohol use |  |  |  |  |  | money saving, restricting access to money or minimal charges for withdrawals. |  |  | of vaginal or anal sex acts where no condom was used among participants who were unmarried or were married and reported extramarital partners<br>Number of unprotected sex acts with casual partners, clients, or CSWs, measured as number of unprotected vaginal or anal sex acts with these types of partners.<br>Sexual risk measured as having ever met a sex partner at a drinking establishment (y/n).<br>AUDIT and AUDASIS<br>measured problematic alcohol use<br>Two questions measured use of commitment savings products | odds of problematic alcohol use | quantified either by frequency or amount. Assessment of commitment savings may have excluded other forms of informal commitment savings. Because of the peculiar characteristics of fishing communities, outcomes can only be generalized to similar settings. The quota-based snowball sampling procedures did not offer equal probability for the selection of participants. |
| Sileo et al., 2021 | The Intersection of Inequitable Gender Norm Endorsement and HIV Stigma: Implications for HIV Care Engagement for Men in Ugandan Fishing Communities | Uganda | 18+ | Male | 300 | Alcohol | Gender-transformative programmes: By addressing detrimental male gender norms, advance gender equality and addressing stigma associated with ART enrolment among men. | Male fisherfolk on ART | Cross-sectional design | Inequitable gender norms measured through the Inequitable Gender Norms (IGN) scale<br>HIV stigma measured using Earnshaw's HIV stigma framework measures<br>First engagement outcome, measured from missed HIV clinic visits from clinic records and self reports. | Retention in HIV care | Cross sectional design limits inference of a causal relationship. The use of self-reported HIV care engagement outcomes is subject to social desirability and recall bias. There was no systematic measure of the degree of agreement between self-report and clinic records. Generalization is limited to male fisherfolk on HIV treatment in Uganda. Likely selection bias which may have influenced the interaction results. |

## Discussion

This is a systematic review on antiretroviral therapy adherence interventions among persons who use AOS in fisherfolk communities. Despite the high prevalence of AOS use in fisherfolk communities and associated barriers to ART adherence, no systematic review has been undertaken in this population to identify and assess the quality of interventions that addresses these barriers and encourage ART adherence. To our knowledge, this is the first review to fill this gap in literature. Interventions aimed at mitigating HIV infections and transmission among persons who use AOS in fisherfolk communities with the possibilities of providing in depth understanding of how peculiarities in this population are being addressed. The review identified five papers from two studies which were included in the analysis thus expanding prior reviews on ART adherence in key and hard to reach populations (Strathdee et al., 2012, Sileo et al., 2019b). Findings from this review highlight several issues on the current state of evidence in this key population. First, there is a paucity of studies in this population in many countries, particularly outside Africa despite the high burden of HIV and alcohol consumption. Second, these interventions have not been tested beyond self-report by the participants and descriptive analysis. However key interventions have been identified, these include counselling, peer support, SBI, economic straightening, social network, gender transformative programmes and prescription monitoring.

Overall, calls have been made on the need to accommodate structural barriers against key and hard to reach populations with the development of models of care that are tailored to specific needs (Macdonald et al., 2017; Sileo et al., 2019c), policies and institution level change that addresses gender inequalities in HIV treatment interventions (Dovel et al., 2015; Fleming and Dworkin, 2016). However, occupational and structural factors specific to fisherfolk communities and risk factors for alcohol use remained as barriers to ART adherence irrespective of motivations for the interventions and seems not to have been adequately addressed in this review. Such risk factors include dealing with cold, wind, long fishing hours, ability to sleep well during off hours and as a motivation to initiate conversation with the opposite sex (Sileo et al., 2019a).

### Strengths and limitations

It is difficult to generalize the findings beyond alcohol consumption. Only 6% of the sample reported using other substances besides alcohol. Only qualitative, and descriptive quantitative studies were found to have met the inclusion criteria. No quantitative randomized or non-randomized control trails have been conducted to investigate any of these interventions in this population. Only studies conducted in English were included, this may have excluded some studies with substantial evidence that were conducted in other languages. Even though the review was not restricted to any country, the included studies were conducted in communities in one country, Uganda in Africa, therefore limits generalization of findings to other countries. In addition, the participants were mainly males (72%) thus limiting generalization of findings to both genders. Another limitation is that none of the studies established whether improved adherence led to better clinical outcomes. One of the previous studies reported that interventions to encourage ART adherence and improved adherence to ART did not translate to improved clinical outcomes (Mbuagbaw et al., 2015). Despite these, this is the first attempt to conduct a systematic review on this key population. The review was a mixed method systematic review that allows for a more comprehensive analysis of ART adherence even with the very limited data. The study therefore provides valuable insight into interventions that have encouraged ART adherence in this key and hard to reach population. Of particular significance is the identification and impacts of new interventions such as the gender transformative programmes and commitment savings.

## Conclusion

Fishing communities are some of the hidden and most-at-risk populations for HIV, AOS use. The physical, structural, cultural and social environment makes interventions to reduce AOS use and encourage ART adherence in this population challenging. Notwithstanding, in this review, a number of interventions were identified. Findings highlight the need for alcohol use reduction interventions for fisherfolk on ART to improve adherence particularly among men. Also added support to the use of gender transformative interventions in altering inequitable gender norms.

The findings highlight some important knowledge gaps that need to be addressed with this population. That the interventions were deduced from non-randomized designs, there is a need for further research aimed at examining the influence of these interventions on ART outcomes in this population and in more countries, develop and test the effectiveness of the interventions in quantitative studies and randomized controlled trails. Considering the universal target of 95%-95%-95% by 2025 and an end to HIV/AIDS scourge by 2030 (UNAIDS, 2014), there is the need to focus attention on key and hard to reach populations with high HIV prevalence and significant barriers to care.

## Data Availability

All data produced in the present work are contained in the manuscript

## References

Abrahams, N., Jewkes, R., Lombard, C., Mathews, S., Campbell, J., & Meel, B. (2010). Impact of telephonic psycho-social support on adherence to post-exposure prophylaxis (PEP) after rape. AIDS care, 22(10), 1173–1181.

Allison, E. H. & Mvula, P. M. (2002). Fishing Livelihoods and Fisheries Management in Malawi. LADDER Working Paper No. 22. https://assets.publishing.service.gov.uk/media/57a08d45e5274a31e000176c/Ladder-wp22.pdf (accessed on 30 April 2023)

Amin, P., & Douaihy, A. (2018). Substance Use Disorders in People Living with Human Immunodeficiency Virus/AIDS. The Nursing clinics of North America, 53(1), 57–65.

Assefa, Y., & Gilks, C. F. (2020). Ending the epidemic of HIV/AIDS by 2030: Will there be an endgame to HIV, or an endemic HIV requiring an integrated health systems response in many countries?. International journal of infectious diseases: IJID: official publication of the International Society for Infectious Diseases, 100, 273–277.

Attonito, J., Villalba, K., & Dévieux, J. G. (2020). Effectiveness of an Intervention for Improving Treatment Adherence, Service Utilization and Viral Load Among HIV-Positive Adult Alcohol Users. AIDS and behavior, 24(5), 1495–1504.

Babor, T. F., McRee, B. G., Kassebaum, P. A., Grimaldi, P. L., Ahmed, K., & Bray, J. (2007). Screening, Brief Intervention, and Referral to Treatment (SBIRT): toward a public health approach to the management of substance abuse. Substance abuse, 28(3), 7–30.

Baker, P., Dworkin, S. L., Tong, S., Banks, I., Shand, T., & Yamey, G. (2014). The men’s health gap: men must be included in the global health equity agenda. Bulletin of the World Health Organization, 92(8), 618–620

Bares, S. H., & Scarsi, K. K. (2022). A new paradigm for antiretroviral delivery: long-acting cabotegravir and rilpivirine for the treatment and prevention of HIV. Current opinion in HIV and AIDS, 17(1), 22–31

Barker, G., & Ricardo, C. (2005). Young Men and the Construction of Masculinity in sub-Saharan Africa: Implications for HIV/AIDS, Conflict, and Violence. Social Development Papers: Conflict Prevention and Reconstruction, Paper 26. Washington, DC: The World Bank.

Bärnighausen, T., Tanser, F., Dabis, F., & Newell, M. L. (2012). Interventions to improve the performance of HIV health systems for treatment-as-prevention in sub-Saharan Africa: the experimental evidence. Current opinion in HIV and AIDS, 7(2), 140–150.

Baron, D., Scorgie, F., Ramskin, L., Khoza, N., Schutzman, J., Stangl, A., Harvey, S., Delany-Moretlwe, S., & EMPOWER study team (2020). “You talk about problems until you feel free”: South African adolescent girls’ and young women’s narratives on the value of HIV prevention peer support clubs. BMC public health, 20(1), 1016.

Bateganya, M. H., Dong, M., Oguntomilade, J., & Suraratdecha, C. (2015). The impact of social services interventions in developing countries: a review of the evidence of impact on clinical outcomes in people living with HIV. Journal of acquired immune deficiency syndromes (1999), *68 Suppl 3*(0 3), S357–S367.

Beckham, S. W., Beyrer, C., Luckow, P., Doherty, M., Negussie, E. K., & Baral, S. D. (2016). Marked sex differences in all-cause mortality on antiretroviral therapy in low- and middle-income countries: a systematic review and meta-analysis. Journal of the International AIDS Society, 19(1), 21106.

Berrien, V. M., Salazar, J. C., Reynolds, E., McKay, K., & HIV Medication Adherence Intervention Group (2004). Adherence to antiretroviral therapy in HIV-infected pediatric patients improves with home-based intensive nursing intervention. AIDS patient care and STDs, 18(6), 355–363.

Bien-Gund, C. H., Ho, J. I., Bair, E. F., Marcus, N., Choi, R. J., Szep, Z., Althoff, A., Momplaisir, F. M., & Thirumurthy, H. (2021). Brief Report: Financial Incentives and Real-Time Adherence Monitoring to Promote Daily Adherence to HIV Treatment and Viral Suppression Among People Living With HIV: A Pilot Study. Journal of acquired immune deficiency syndromes (1999), 87(1), 688–692.

Bogart, L. M., Matovu, J. K. B., Wagner, G. J., Green, H. D., Storholm, E. D., Klein, D. J., Marsh, T., MacCarthy, S., & Kambugu, A. (2020). A Pilot Test of Game Changers, a Social Network Intervention to Empower People with HIV to be Prevention Advocates in Uganda. AIDS and behavior, 24(9), 2490–2508.

Bogart, L. M., Naigino, R., Maistrellis, E., Wagner, G. J., Musoke, W., Mukasa, B., Jumamil, R., & Wanyenze, R. K. (2016). Barriers to Linkage to HIV Care in Ugandan Fisherfolk Communities: A Qualitative Analysis. AIDS and behavior, 20(10), 2464–2476.

Bonnevie, E., Kigozi, G., Kairania, R., Ssemanda, J. B., Nakyanjo, N., Ddaaki, W. G., Ssekyewa, C., & Wagman, J. A. (2020). Alcohol use in fishing communities and men’s willingness to participate in an alcohol, violence and HIV risk reduction intervention: qualitative findings from Rakai, Uganda. Culture, health & sexuality, 22(3), 275–291.

Brown, M. J., & Adeagbo, O. (2022). Trauma-Informed HIV Care Interventions: Towards a Holistic Approach. Current HIV/AIDS reports, 19(3), 177–183.

Brown, T., & George, P. (2019). Perceptions of opioid misuse and chronic pain: A qualitative assessment of Rhode Island commercial fishing captains. Journal of opioid management, 15(2), 129–135.

Brown, S. E., Wickersham, J. A., Pelletier, A. R., Marcus, R. M., Erenrich, R., Kamarulzaman, A., & Altice, F. L. (2017). Attitudes toward medication-assisted treatment among fishermen in Kuantan, Malaysia, who inject drugs. Journal of ethnicity in substance abuse, 16(3), 363–379.

Burke, V. M., Nakyanjo, N., Ddaaki, W., Payne, C., Hutchinson, N., Wawer, M. J., Nalugoda, F., & Kennedy, C. E. (2017). HIV self-testing values and preferences among sex workers, fishermen, and mainland community members in Rakai, Uganda: A qualitative study. PloS one, 12(8), e0183280.

Burgos-Soto, J., Ben Farhat, J., Alley, I., Ojuka, P., Mulogo, E., Kise-Sete, T., Bouhenia, M., Salumu, L., Mathela, R., Langendorf, C., Cohuet, S., & Huerga, H. (2020). HIV epidemic and cascade of care in 12 east African rural fishing communities: results from a population-based survey in Uganda. BMC public health, 20(1), 970.

Byron, E., Gillespie, S., & Nangami, M. (2008). Integrating nutrition security with treatment of people living with HIV: lessons from Kenya. Food and nutrition bulletin, 29(2), 87–97.

Carbone, N. B., Njala, J., Jackson, D. J., Eliya, M. T., Chilangwa, C., Tseka, J., Zulu, T., Chinkonde, J. R., Sherman, J., Zimba, C., Mofolo, I. A., & Herce, M. E. (2019). “I would love if there was a young woman to encourage us, to ease our anxiety which we would have if we were alone”: Adapting the Mothers2Mothers Mentor Mother Model for adolescent mothers living with HIV in Malawi. PloS one, 14(6), e0217693.

Carey, M. P., Dunne, E. M., Norris, A., Dunsiger, S., Rich, C., Rosen, R. K., Chan, P., & Salmoirago-Blotcher, E. (2020). Telephone-Delivered Mindfulness Training to Promote Medication Adherence and Reduce Sexual Risk Behavior Among Persons Living with HIV: An Exploratory Clinical Trial. AIDS and behavior, 24(6), 1912–1928.

Carpenter, C. C., Fischl, M. A., Hammer, S. M., Hirsch, M. S., Jacobsen, D. M., Katzenstein, D. A., Montaner, J. S., Richman, D. D., Sáag, M. S., Schooley, R. T., Thompson, M. A., Vella, S., Yeni, P. G., & Volberding, P. A. (1996). Antiretroviral therapy for HIV infection in 1996. Recommendations of an international panel. International AIDS Society-USA. JAMA, 276(2), 146–154.

CDC (January 26, 2023). Commercial fishing safety. https://www.cdc.gov/niosh/topics/fishing (accessed on 12 April 2023).

CDC (2018). HIV surveillance report, 2017; vol 29. http://www.cdc.gov/hiv/library/reports/hiv-surveillance.html. Published November 2018 (accessed on 2 April 2023)

Chaiyachati, K. H., Ogbuoji, O., Price, M., Suthar, A. B., Negussie, E. K., & Bärnighausen, T. (2014). Interventions to improve adherence to antiretroviral therapy: a rapid systematic review. *AIDS (London*, England), 28 *Suppl 2*, S187–S204.

Chang, L. W., Grabowski, M. K., Ssekubugu, R., Nalugoda, F., Kigozi, G., Nantume, B., Lessler, J., Moore, S. M., Quinn, T. C., Reynolds, S. J., Gray, R. H., Serwadda, D., & Wawer, M. J. (2016). Heterogeneity of the HIV epidemic in agrarian, trading, and fishing communities in Rakai, Uganda: an observational epidemiological study. The lancet. HIV, 3(8), e388–e396.

Chesney, M. A., Ickovics, J. R., Chambers, D. B., Gifford, A. L., Neidig, J., Zwickl, B., & Wu, A. W. (2000). Self-reported adherence to antiretroviral medications among participants in HIV clinical trials: the AACTG adherence instruments. Patient Care Committee & Adherence Working Group of the Outcomes Committee of the Adult AIDS Clinical Trials Group (AACTG). AIDS care, 12(3), 255–266.

Chikovore, J., Gillespie, N., McGrath, N., Orne-Gliemann, J., Zuma, T., & ANRS 12249 TasP Study Group (2016). Men, masculinity, and engagement with treatment as prevention in KwaZulu-Natal, South Africa. AIDS care, 28 Suppl 3(Suppl 3), 74–82.

Choo, M. K., El-Bassel, N., Adam, P. C., Gilbert, L., Wu, E., West, B. S., Bazazi, A. R., De Wit, J. B., Ismail, R., & Kamarulzaman, A. (2015). Prevalence and Correlates of HIV and Hepatitis C Virus Infections and Risk Behaviors among Malaysian Fishermen. PloS one, 10(8), e0118422.

Chung, M. H., Richardson, B. A., Tapia, K., Benki-Nugent, S., Kiarie, J. N., Simoni, J. M., Overbaugh, J., Attwa, M., & John-Stewart, G. C. (2011). A randomized controlled trial comparing the effects of counseling and alarm device on HAART adherence and virologic outcomes. PLoS medicine, 8(3), e1000422.

Collier, A. C., Ribaudo, H., Mukherjee, A. L., Feinberg, J., Fischl, M. A., Chesney, M., & Adult AIDS Clinical Trials Group 746 Substudy Team (2005). A randomized study of serial telephone call support to increase adherence and thereby improve virologic outcome in persons initiating antiretroviral therapy. The Journal of infectious diseases, 192(8), 1398–1406.

Cooper, V., Horne, R., Gellaitry, G., Vrijens, B., Lange, A. C., Fisher, M., & White, D. (2010). The impact of once-nightly versus twice-daily dosing and baseline beliefs about HAART on adherence to efavirenz-based HAART over 48 weeks: the NOCTE study. Journal of acquired immune deficiency syndromes (1999), *53*(3), 369–377.

Cornell, M., McIntyre, J., & Myer, L. (2011). Men and antiretroviral therapy in Africa: our blind spot. Tropical medicine & international health: TM & IH, 16(7), 828–829.

De Boni, R., Veloso, V. G., & Grinsztejn, B. (2014). Epidemiology of HIV in Latin America and the Caribbean. Current opinion in HIV and AIDS, 9(2), 192–198.

Dejesus, E., Young, B., Morales-Ramirez, J. O., Sloan, L., Ward, D. J., Flaherty, J. F., Ebrahimi, R., Maa, J. F., Reilly, K., Ecker, J., McColl, D., Seekins, D., Farajallah, A., & AI266073 Study Group (2009). Simplification of antiretroviral therapy to a single-tablet regimen consisting of efavirenz, emtricitabine, and tenofovir disoproxil fumarate versus unmodified antiretroviral therapy in virologically suppressed HIV-1-infected patients. Journal of acquired immune deficiency syndromes (1999), 51(2), 163–174.

Derbalah, A., Karpick, H. C., Maize, H., Skersick, P., Cottrell, M., & Rao, G. G. (2022). Role of islatravir in HIV treatment and prevention: an update. Current opinion in HIV and AIDS, 17(4), 240–246.

Deryabina, A. P., & El-Sadr, W. M. (2019). Optimizing HIV prevention and treatment outcomes for persons with substance use in Central Asia: what will it take?. Current opinion in HIV and AIDS, 14(5), 374–380.

Detsis, M., Tsioutis, C., Karageorgos, S. A., Sideroglou, T., Hatzakis, A., & Mylonakis, E. (2017). Factors Associated with HIV Testing and HIV Treatment Adherence: A Systematic Review. Current pharmaceutical design, 23(18), 2568–2578.

DiIorio, C., McCarty, F., Resnicow, K., McDonnell Holstad, M., Soet, J., Yeager, K., Sharma, S. M., Morisky, D. E., & Lundberg, B. (2008). Using motivational interviewing to promote adherence to antiretroviral medications: a randomized controlled study. AIDS care, 20(3), 273– 283.

Dovel, K., Yeatman, S., Watkins, S., & Poulin, M. (2015). Men’s heightened risk of AIDS-related death: the legacy of gendered HIV testing and treatment strategies. *AIDS (London*, England), 29(10), 1123–1125.

Duncan, L. G., Moskowitz, J. T., Neilands, T. B., Dilworth, S. E., Hecht, F. M., & Johnson, M. O. (2012). Mindfulness-based stress reduction for HIV treatment side effects: a randomized, wait-list controlled trial. Journal of pain and symptom management, 43(2), 161–171.

Dupas, P., & Robinson, J. (2013). Why Don’t the Poor Save More? Evidence from Health Savings Experiments. The American economic review, 103(4), 1138–1171.

El-Bassel, N., Shaw, S. A., Dasgupta, A., & Strathdee, S. A. (2014). Drug use as a driver of HIV risks: re-emerging and emerging issues. Current opinion in HIV and AIDS, 9(2), 150–155.

Enane, L. A., Apondi, E., Aluoch, J., Bakoyannis, G., Lewis Kulzer, J., Kwena, Z., Kantor, R., Chory, A., Gardner, A., Scanlon, M., Goodrich, S., Wools-Kaloustian, K., Elul, B., & Vreeman, R. C. (2021). Social, economic, and health effects of the COVID-19 pandemic on adolescents retained in or recently disengaged from HIV care in Kenya. PloS one, 16(9), e0257210.

Essex, M., Makhema, J., & Lockman, S. (2019). Reaching 90-90-90 in Botswana. Current opinion in HIV and AIDS, 14(6), 442–448.

Ferretti, F., & Boffito, M. (2018). Rilpivirine long-acting for the prevention and treatment of HIV infection. Current opinion in HIV and AIDS, 13(4), 300–307.

Figueroa, M. E., Poppe, P., Carrasco, M., Pinho, M. D., Massingue, F., Tanque, M., & Kwizera, A. (2016). Effectiveness of Community Dialogue in Changing Gender and Sexual Norms for HIV Prevention: Evaluation of the Tchova Tchova Program in Mozambique. Journal of health communication, 21(5), 554–563.

Finitsis, D. J., Pellowski, J. A., & Johnson, B. T. (2014). Text message intervention designs to promote adherence to antiretroviral therapy (ART): a meta-analysis of randomized controlled trials. PloS one, 9(2), e88166.

Fisher, J. D., Amico, K. R., Fisher, W. A., Cornman, D. H., Shuper, P. A., Trayling, C., Redding, C., Barta, W., Lemieux, A. F., Altice, F. L., Dieckhaus, K., Friedland, G., & LifeWindows Team (2011). Computer-based intervention in HIV clinical care setting improves antiretroviral adherence: the LifeWindows Project. AIDS and behavior, 15(8), 1635–1646.

Fleming, P. J., & Dworkin, S. L. (2016). The importance of masculinity and gender norms for understanding institutional responses to HIV testing and treatment strategies. *AIDS (London*, England), 30(1), 157–158.

Ford, N., Nachega, J. B., Engel, M. E., & Mills, E. J. (2009). Directly observed antiretroviral therapy: a systematic review and meta-analysis of randomised clinical trials. *Lancet (London*, England), 374(9707), 2064–2071.

Ford, K., & Chamratrithirong, A. (2008). Migrant seafarers and HIV risk in Thai communities. AIDS education and prevention : official publication of the International Society for AIDS Education, 20(5), 454–463.

Fort, E., Massardier-Pilonchéry, A., Facy, F., & Bergeret, A. (2012). Prevalence of drug use in French seamen. Addictive behaviors, 37(3), 335–338.

Fuge, T. G., Tsourtos, G., & Miller, E. R. (2022). Factors affecting optimal adherence to antiretroviral therapy and viral suppression amongst HIV-infected prisoners in South Ethiopia: a comparative cross-sectional study. AIDS research and therapy, 19(1), 5.

Gottert, A. (2014). Gender norms, masculine gender-role strain, and HIV risk behaviors among men in rural South Africa. https://www.semanticscholar.org/paper/Gender-norms%2C-masculine-gender-role-strain%2C-and-HIV-Gottert/0d9f8c471cdf1983130ed74961a46e72021ddc0c#citing-papers (accessed on 10 April 2023).

Grimsrud, A., Wilkinson, L., Eshun-Wilson, I., Holmes, C., Sikazwe, I., & Katz, I. T. (2020). Understanding Engagement in HIV Programmes: How Health Services Can Adapt to Ensure No One Is Left Behind. Current HIV/AIDS reports, 17(5), 458–466.

Hardy, H., Kumar, V., Doros, G., Farmer, E., Drainoni, M. L., Rybin, D., Myung, D., Jackson, J., Backman, E., Stanic, A., & Skolnik, P. R. (2011). Randomized controlled trial of a personalized cellular phone reminder system to enhance adherence to antiretroviral therapy. AIDS patient care and STDs, 25(3), 153–161.

Hendershot, C. S., Stoner, S. A., Pantalone, D. W., & Simoni, J. M. (2009). Alcohol use and antiretroviral adherence: review and meta-analysis. Journal of acquired immune deficiency syndromes (1999), 52(2), 180–202.

Hickey, M. D., Salmen, C. R., Omollo, D., Mattah, B., Fiorella, K. J., Geng, E. H., Bacchetti, P., Blat, C., Ouma, G. B., Zoughbie, D., Tessler, R. A., Salmen, M. R., Campbell, H., Gandhi, M., Shade, S., Njoroge, B., Bukusi, E. A., & Cohen, C. R. (2015). Implementation and Operational Research: Pulling the Network Together: Quasiexperimental Trial of a Patient-Defined Support Network Intervention for Promoting Engagement in HIV Care and Medication Adherence on Mfangano Island, Kenya. Journal of acquired immune deficiency syndromes (1999), 69(4), e127–e134.

Hill, S., & Kavookjian, J. (2012). Motivational interviewing as a behavioral intervention to increase HAART adherence in patients who are HIV-positive: a systematic review of the literature. AIDS care, 24(5), 583–592.

HIV Prevention Trials Network (2016). Publication of HPTN 052 Final Results: HIV Treatment Offers Durable Prevention of HIV Transmission in Sero-discordant Couples. The HIV Prevention Trials Network. Available from https://www.hptn.org/news-and-events/press-releases/publication-of-hptn-052-final-results-hiv-treatment-offers-durable (accessed on 1 April 2023).

Hlophe, L. D., Tamuzi, J. L., Shumba, C. S., & Nyasulu, P. S. (2023). Barriers and facilitators to anti-retroviral therapy adherence among adolescents aged 10 to 19 years living with HIV in sub-Saharan Africa: A mixed-methods systematic review and meta-analysis. PloS one, 18(5), e0276411.

Holstad, M. M., Essien, J. E., Ekong, E., Higgins, M., Teplinskiy, I., & Adewuyi, M. F. (2012). Motivational groups support adherence to antiretroviral therapy and use of risk reduction behaviors in HIV positive Nigerian women: a pilot study. African journal of reproductive health, 16(3), 14–27.

Holstad, M. M., DiIorio, C., Kelley, M. E., Resnicow, K., & Sharma, S. (2011). Group motivational interviewing to promote adherence to antiretroviral medications and risk reduction behaviors in HIV infected women. AIDS and behavior, 15(5), 885–896.

Holtgrave D. R. (2005). Causes of the decline in AIDS deaths, United States, 1995-2002: prevention, treatment or both?. International journal of STD & AIDS, 16(12), 777–781.

Hong, Q. N., Pluyi, P., Fabregues, S., Bartlett, G., Boardman, F., Cargo, M., Dagenais, P., Gagnon, M. P., Griffiths, F., Nicolau, B., O’Cathain, A., Rousseau., M. C., Vedel, L. (2018). Mixed Methods Appraisal Tool (MMAT), version 2018. Registration of Copyright (#1148552), Canadian Intellectual Property Office, Industry Canada.

Joseph, B., Kerr, T., Puskas, C. M., Montaner, J., Wood, E., & Milloy, M. J. (2015). Factors linked to transitions in adherence to antiretroviral therapy among HIV-infected illicit drug users in a Canadian setting. AIDS care, 27(9), 1128–1136.

Justice, A. C., McGinnis, K. A., Tate, J. P., Braithwaite, R. S., Bryant, K. J., Cook, R. L., Edelman, E. J., Fiellin, L. E., Freiberg, M. S., Gordon, A. J., Kraemer, K. L., Marshall, B. D., Williams, E. C., & Fiellin, D. A. (2016). Risk of mortality and physiologic injury evident with lower alcohol exposure among HIV infected compared with uninfected men. Drug and alcohol dependence, 161, 95–103.

Kagaayi, J., Chang, L. W., Ssempijja, V., Grabowski, M. K., Ssekubugu, R., Nakigozi, G., Kigozi, G., Serwadda, D. M., Gray, R. H., Nalugoda, F., Sewankambo, N. K., Nelson, L., Mills, L. A., Kabatesi, D., Alamo, S., Kennedy, C. E., Tobian, A. A. R., Santelli, J. S., Ekström, A. M., Nordenstedt, H., … Reynolds, S. J. (2019). Impact of combination HIV interventions on HIV incidence in hyperendemic fishing communities in Uganda: a prospective cohort study. The lancet. HIV, 6(10), e680–e687.

Kagee, A., Swartz, A., & Swartz, L. (2014). Theorising beyond the individual: adherence to antiretroviral therapy in resource-constrained societies. Journal of health psychology, 19(1), 103–109.

Kalichman, S. C., Cherry, C., Kalichman, M. O., Amaral, C. M., White, D., Pope, H., Swetzes, C., Eaton, L., Macy, R., & Cain, D. (2011). Integrated behavioral intervention to improve HIV/AIDS treatment adherence and reduce HIV transmission. American journal of public health, 101(3), 531–538.

Kaner, E. F., Beyer, F., Dickinson, H. O., Pienaar, E., Campbell, F., Schlesinger, C., Heather, N., Saunders, J., & Burnand, B. (2007). Effectiveness of brief alcohol interventions in primary care populations. The Cochrane database of systematic reviews, (2), CD004148.

Kanters, S., Park, J. J., Chan, K., Ford, N., Forrest, J., Thorlund, K., Nachega, J. B., & Mills, E. J. (2016). Use of peers to improve adherence to antiretroviral therapy: a global network meta-analysis. Journal of the International AIDS Society, 19(1), 21141.

Kapesa, A., Basinda, N., Nyanza, E. C., Mushi, M. F., Jahanpour, O., & Ngallaba, S. E. (2018). Prevalence of HIV infection and uptake of HIV/AIDS services among fisherfolk in landing Islands of Lake Victoria, north western Tanzania. BMC health services research, 18(1), 980.

Keyes, K. M., Hatzenbuehler, M. L., & Hasin, D. S. (2011). Stressful life experiences, alcohol consumption, and alcohol use disorders: the epidemiologic evidence for four main types of stressors. Psychopharmacology, 218(1), 1–17.

Kiene, S. M., Ediau, M., Schmarje, K. A., Kintu, M., & Tumwesigye, N. M. (2019a). Exploring the Potential of Savings-Led Economic Strengthening HIV Interventions Among High-Risk Economically Vulnerable Fishing Communities in Uganda: Associations Between Use of Commitment Savings, Sexual Risk Behavior, and Problematic Alcohol Use. AIDS and behavior, 23(9), 2347–2360.

Kiene, S. M., Sileo, K. M., Dove, M., & Kintu, M. (2019b). Hazardous alcohol consumption and alcohol-related problems are associated with unknown and HIV-positive status in fishing communities in Uganda. AIDS care, 31(4), 451–459.

Kiene, S. M., Simbayi, L. C., Abrams, A., Cloete, A., Tennen, H., & Fisher, J. D. (2008). High rates of unprotected sex occurring among HIV-positive individuals in a daily diary study in South Africa: the role of alcohol use. Journal of acquired immune deficiency syndromes (1999), *49*(2), 219–226.

Kilburn, K. N., Pettifor, A., Edwards, J. K., Selin, A., Twine, R., MacPhail, C., Wagner, R., Hughes, J. P., Wang, J., & Kahn, K. (2018). Conditional cash transfers and the reduction in partner violence for young women: an investigation of causal pathways using evidence from a randomized experiment in South Africa (HPTN 068). Journal of the International AIDS Society, 21 Suppl 1(Suppl Suppl 1), e25043.

Kissling, E., Allison, E. H., Seeley, J. A., Russell, S., Bachmann, M., Musgrave, S. D., & Heck, S. (2005). Fisherfolk are among groups most at risk of HIV: cross-country analysis of prevalence and numbers infected. *AIDS (London*, England), 19(17), 1939–1946.

Kuteesa, M. O., Webb, E. L., Kawuma, R., Naluwugge, J., Thadeus, K., Ndekezi, D. & Seeley, J. (2022). ’We shall drink until Lake Victoria dries up’: Drivers of heavy drinking and illicit drug use among young Ugandans in fishing communities. Global public health, 17(4), 538–554.

Kuteesa, M. O., Weiss, H. A., Cook, S., Seeley, J., Ssentongo, J. N., Kizindo, R., Ngonzi, P., Sewankambo, M., & Webb, E. L. (2020). Epidemiology of Alcohol Misuse and Illicit Drug Use Among Young People Aged 15-24 Years in Fishing Communities in Uganda. International journal of environmental research and public health, 17(7), 2401.

Ky S., Rugemintwari, C. & Sauviat, A. (2016). Does mobile money affect saving behavior? Evidence from a developing country. https://papers.ssrn.com/sol3/papers.cfm?abstract_id=2815090#:~:text=Our%20main%20results%20show%20that%2C%20although%20using%20mobile,propensity%20of%20individuals%20to%20save%20for%20health%20emergencies (accessed on 12 April 2023).

Lappalainen, L., Nolan, S., Dobrer, S., Puscas, C., Montaner, J., Ahamad, K., Dong, H., Kerr, T., Wood, E., & Milloy, M. J. (2015). Dose-response relationship between methadone dose and adherence to antiretroviral therapy among HIV-positive people who use illicit opioids. Addiction (Abingdon, England), 110(8), 1330–1339.

Laraqui, O., Laraqui, S., Manar, N., Ghailan, T., Deschamps, F., & Laraqui, C. H. (2017). Prevalence of consumption of addictive substances amongst Moroccan fishermen. International maritime health, 68(1), 19–25.

Leeman, J., Chang, Y. K., Lee, E. J., Voils, C. I., Crandell, J., & Sandelowski, M. (2010). Implementation of antiretroviral therapy adherence interventions: a realist synthesis of evidence. Journal of advanced nursing, 66(9), 1915–1930.

Lester, R. T., Ritvo, P., Mills, E. J., Kariri, A., Karanja, S., Chung, M. H., Jack, W., Habyarimana, J., Sadatsafavi, M., Najafzadeh, M., Marra, C. A., Estambale, B., Ngugi, E., Ball, T. B., Thabane, L., Gelmon, L. J., Kimani, J., Ackers, M., & Plummer, F. A. (2010). Effects of a mobile phone short message service on antiretroviral treatment adherence in Kenya (WelTel Kenya1): a randomised trial. *Lancet (London*, England), 376(9755), 1838–1845.

Lockman, S., & Sax, P. (2012). Treatment-for-prevention: clinical considerations. Current opinion in HIV and AIDS, 7(2), 131–139.

Long, A., Mbabali, I., Hutton, H. E., Thomas, A. G., Bugos, E., Mulamba, J., Amico, K. R., Nalugoda, F., Gray, R. H., Wawer, M. J., Nakigozi, G., & Chang, L. W. (2017). Design and Implementation of a Community Health Worker HIV Treatment and Prevention Intervention in an HIV Hot Spot Fishing Community in Rakai, Uganda. Journal of the International Association of Providers of AIDS Care, 16(5), 499–505.

Lubega, M., Nakyaanjo, N., Nansubuga, S., Hiire, E., Kigozi, G., Nakigozi, G., Lutalo, T., Nalugoda, F., Serwadda, D., Gray, R., Wawer, M., Kennedy, C., & Reynolds, S. J. (2015). Understanding the socio-structural context of high HIV transmission in kasensero fishing community, South Western Uganda. BMC public health, 15, 1033.

Macdonald, V., Verster, A., & Baggaley, R. (2017). A call for differentiated approaches to delivering HIV services to key populations. Journal of the International AIDS Society, 20(Suppl 4), 21658.

MacPherson, E. E., Sadalaki, J., Njoloma, M., Nyongopa, V., Nkhwazi, L., Mwapasa, V., Lalloo, D. G., Desmond, N., Seeley, J., & Theobald, S. (2012). Transactional sex and HIV: understanding the gendered structural drivers of HIV in fishing communities in Southern Malawi. Journal of the International AIDS Society, 15 Suppl 1(Suppl 1), 1–9.

Mafigiri, R., Matovu, J. K., Makumbi, F. E., Ndyanabo, A., Nabukalu, D., Sakor, M., Kigozi, G., Nalugoda, F., & Wanyenze, R. K. (2017). HIV prevalence and uptake of HIV/AIDS services among youths (15-24 Years) in fishing and neighboring communities of Kasensero, Rakai District, South Western Uganda. BMC public health, 17(1), 251.

Maitland, D., Jackson, A., Osorio, J., Mandalia, S., Gazzard, B. G., Moyle, G. J., & Epivir-Ziagen (EZ) Switch Study Team (2008). Switching from twice-daily abacavir and lamivudine to the once-daily fixed-dose combination tablet of abacavir and lamivudine improves patient adherence and satisfaction with therapy. HIV medicine, 9(8), 667–672.

Mannheimer, S., & Hirsch-Moverman, Y. (2015). What we know and what we do not know about factors associated with and interventions to promote antiretroviral adherence. Current infectious disease reports, 17(4), 466.

Marcus, J. L., Chao, C. R., Leyden, W. A., Xu, L., Quesenberry, C. P., Jr, Klein, D. B., Towner, W. J., Horberg, M. A., & Silverberg, M. J. (2016). Narrowing the Gap in Life Expectancy Between HIV-Infected and HIV-Uninfected Individuals With Access to Care. Journal of acquired immune deficiency syndromes (1999), 73(1), 39–46.

Mbuagbaw, L., Sivaramalingam, B., Navarro, T., Hobson, N., Keepanasseril, A., Wilczynski, N. J., Haynes, R. B., & Patient Adherence Review (PAR) Team (2015). Interventions for Enhancing Adherence to Antiretroviral Therapy (ART): A Systematic Review of High Quality Studies. AIDS patient care and STDs, 29(5), 248–266.

McManus, H., O’Connor, C. C., Boyd, M., Broom, J., Russell, D., Watson, K., Roth, N., Read, P. J., Petoumenos, K., Law, M. G., & Australian HIV Observational Database (2012). Long-term survival in HIV positive patients with up to 15 Years of antiretroviral therapy. PloS one, 7(11), e48839.

Mendelsohn, J. B., Schilperoord, M., Spiegel, P., & Ross, D. A. (2012). Adherence to antiretroviral therapy and treatment outcomes among conflict-affected and forcibly displaced populations: a systematic review. Conflict and health, 6(1), 9.

Mgabo, M. R., Sospeter, B. M., Buruna, M. I. (2013). Social context and modeling strategies for controlling the spread of HIV/AIDS infections in fishing communities of Lake Victoria. A case of Lukuba Island in Tanzania. Int J Behav Soc Mov Sci. 2(2) 101–17.

Michalopoulos, L. M., Jiwatram-Negrón, T., Choo, M. K., Kamarulzaman, A., & El-Bassel, N. (2016). The association between psychosocial and structural-level stressors and HIV injection drug risk behavior among Malaysian fishermen: A cross-sectional study. BMC public health, 16, 464.

Mills, E. J., Lester, R., Thorlund, K., Lorenzi, M., Muldoon, K., Kanters, S., Linnemayr, S., Gross, R., Calderon, Y., Amico, K. R., Thirumurthy, H., Pearson, C., Remien, R. H., Mbuagbaw, L., Thabane, L., Chung, M. H., Wilson, I. B., Liu, A., Uthman, O. A., Simoni, J., … Nachega, J. B. (2014). Interventions to promote adherence to antiretroviral therapy in Africa: a network meta-analysis. The lancet. HIV, 1(3), e104–e111.

Mohd Salleh, N. A., Richardson, L., Kerr, T., Shoveller, J., Montaner, J., Kamarulzaman, A., & Milloy, M. J. (2018). A Longitudinal Analysis of Daily Pill Burden and Likelihood of Optimal Adherence to Antiretroviral Therapy Among People Living With HIV Who Use Drugs. Journal of addiction medicine, 12(4), 308–314.

Montaner, J. S., Lima, V. D., Harrigan, P. R., Lourenço, L., Yip, B., Nosyk, B., Wood, E., Kerr, T., Shannon, K., Moore, D., Hogg, R. S., Barrios, R., Gilbert, M., Krajden, M., Gustafson, R., Daly, P., & Kendall, P. (2014). Expansion of HAART coverage is associated with sustained decreases in HIV/AIDS morbidity, mortality and HIV transmission: the “HIV Treatment as Prevention” experience in a Canadian setting. PloS one, 9(2), e87872.

Musumari, P. M., Techasrivichien, T., Srithanaviboonchai, K., Wanyenze, R. K., Matovu, J. K. B., Poudyal, H., Suguimoto, S. P., Zamani, S., Tangmunkongvorakul, A., Ono-Kihara, M., & Kihara, M. (2021). HIV epidemic in fishing communities in Uganda: A scoping review. PloS one, 16(4), e0249465.

Naar-King, S., Outlaw, A. Y., Sarr, M., Parsons, J. T., Belzer, M., Macdonell, K., Tanney, M., Ondersma, S. J., & Adolescent Medicine Network for HIV/AIDS Interventions (2013). Motivational Enhancement System for Adherence (MESA): pilot randomized trial of a brief computer-delivered prevention intervention for youth initiating antiretroviral treatment. Journal of pediatric psychology, 38(6), 638–648.

Naigino, R., Miller, A. P., Ediau, M., Anecho, A., Senoga, U., Tumwesigye, N. M., Wanyenze, R. K., Mukasa, B., Hahn, J. A., Reed, E., Sileo, K. M., & Kiene, S. M. (2023). Stakeholder perspectives on the Kisoboka intervention: A behavioral and structural intervention to reduce hazardous alcohol use and improve HIV care engagement among men living with HIV in Ugandan fishing communities. Drug and alcohol dependence, 253, 111011.

Nanavati, J. (2019). Databases Beyond PubMed. https://vdocument.in/download/databases-beyond-pubmed-to-pubmed-medline-a-locate-dissertations-and-book-chapters.html (accessed on 28 April 2023)

Naugle, D. A., Tibbels, N. J., Hendrickson, Z. M., Dosso, A., Van Lith, L., Mallalieu, E. C., Kouadio, A. M., Kra, W., Kamara, D., Dailly-Ajavon, P., Cissé, A., Seifert-Ahanda, K., Thaddeus, S., Babalola, S., & Hoffmann, C. J. (2019). Bringing fear into focus: The intersections of HIV and masculine gender norms in Côte d’Ivoire. PloS one, 14(10), e0223414.

Ndekha, M., van Oosterhout, J. J., Saloojee, H., Pettifor, J., & Manary, M. (2009). Nutritional status of Malawian adults on antiretroviral therapy 1 year after supplementary feeding in the first 3 months of therapy. Tropical medicine & international health: TM & IH, 14(9), 1059–1063.

O’Halloran Leach, E., Lu, H., Caballero, J., Thomas, J. E., Spencer, E. C., & Cook, R. L. (2021). Defining the optimal cut-point of self-reported ART adherence to achieve viral suppression in the era of contemporary HIV therapy: a cross-sectional study. AIDS research and therapy, 18(1), 36.

Okoror, T. A., Falade, C. O., Walker, E. M., Olorunlana, A., & Anaele, A. (2016). Social context surrounding HIV diagnosis and construction of masculinity: a qualitative study of stigma experiences of heterosexual HIV positive men in southwest Nigeria. BMC public health, 16, 507.

Ombere, S. O., & Nyambedha, E. O. (2023). Non-adherence to antiretroviral treatment among migrating fishermen in western Kenya’s islands: a rapid qualitative study. African journal of AIDS research: AJAR, 22(3), 237–243.

Orth, Z., & van Wyk, B. (2021). A Facility-based Family Support Intervention to Improve Treatment Outcomes for Adolescents on Antiretroviral Therapy in the Cape Metropole, South Africa. Journal of the International Association of Providers of AIDS Care, 20, 23259582211059289.

Ouansafi, I., Chibanda, D., Munetsi, E., & Simms, V. (2021). Impact of Friendship Bench problem-solving therapy on adherence to ART in young people living with HIV in Zimbabwe: A qualitative study. PloS one, 16(4), e0250074.

Ousley, J., Nesbitt, R., Kyaw, N. T. T., Bermudez, E., Soe, K. P., Anicete, R., Mon, P. E., Le Shwe Sin Ei, W., Christofani, S., Fernandez, M., & Ciglenecki, I. (2018). Increased hepatitis C virus co-infection and injection drug use in HIV-infected fishermen in Myanmar. BMC infectious diseases, 18(1), 657.

Pantelic, M., Steinert, J. I., Park, J., Mellors, S., & Murau, F. (2019). ’Management of a spoiled identity’: systematic review of interventions to address self-stigma among people living with and affected by HIV. BMJ global health, 4(2), e001285.

Parashar, S., Collins, A. B., Montaner, J. S., Hogg, R. S., & Milloy, M. J. (2016). Reducing rates of preventable HIV/AIDS-associated mortality among people living with HIV who inject drugs. Current opinion in HIV and AIDS, 11(5), 507–513.

Parienti, J. J., Massari, V., Reliquet, V., Chaillot, F., Le Moal, G., Arvieux, C., Vabret, A., Verdon, R., & POSOVIR Study Group (2007). Effect of twice-daily nevirapine on adherence in HIV-1-infected patients: a randomized controlled study. *AIDS (London*, England), 21(16), 2217– 2222.

Parsons, J. T., Golub, S. A., Rosof, E., & Holder, C. (2007). Motivational interviewing and cognitive-behavioral intervention to improve HIV medication adherence among hazardous drinkers: a randomized controlled trial. Journal of acquired immune deficiency syndromes (1999), 46(4), 443–450.

Pearson, C. R., Micek, M. A., Simoni, J. M., Hoff, P. D., Matediana, E., Martin, D. P., & Gloyd, S. S. (2007). Randomized control trial of peer-delivered, modified directly observed therapy for HAART in Mozambique. Journal of acquired immune deficiency syndromes (1999), 46(2), 238–244.

Percin, F., Akyol, O., Davas, A., & Saygi, H. (2012). Occupational health of Turkish Aegean small-scale fishermen. Occupational medicine (Oxford, England), 62(2), 148–151.

Petoumenos, K., & Law, M. G. (2016). Smoking, alcohol and illicit drug use effects on survival in HIV-positive persons. Current opinion in HIV and AIDS, 11(5), 514–520.

Phanuphak, N., & Gulick, R. M. (2020). HIV treatment and prevention 2019: current standards of care. Current opinion in HIV and AIDS, 15(1), 4–12.

Plankey, M. W., Ostrow, D. G., Stall, R., Cox, C., Li, X., Peck, J. A., & Jacobson, L. P. (2007). The relationship between methamphetamine and popper use and risk of HIV seroconversion in the multicenter AIDS cohort study. Journal of acquired immune deficiency syndromes (1999), 45(1), 85–92.

Pop-Eleches, C., Thirumurthy, H., Habyarimana, J. P., Zivin, J. G., Goldstein, M. P., de Walque, D., MacKeen, L., Haberer, J., Kimaiyo, S., Sidle, J., Ngare, D., & Bangsberg, D. R. (2011). Mobile phone technologies improve adherence to antiretroviral treatment in a resource-limited setting: a randomized controlled trial of text message reminders. AIDS (London, England), 25(6), 825–834.

Portsmouth, S. D., Osorio, J., McCormick, K., Gazzard, B. G., & Moyle, G. J. (2005). Better maintained adherence on switching from twice-daily to once-daily therapy for HIV: a 24-week randomized trial of treatment simplification using stavudine prolonged-release capsules. HIV medicine, 6(3), 185–190.

Pradier, C., Bentz, L., Spire, B., Tourette-Turgis, C., Morin, M., Souville, M., Rebillon, M., Fuzibet, J. G., Pesce, A., Dellamonica, P., & Moatti, J. P. (2003). Efficacy of an educational and counseling intervention on adherence to highly active antiretroviral therapy: French prospective controlled study. HIV clinical trials, 4(2), 121–131.

Purcell, D. W., Latka, M. H., Metsch, L. R., Latkin, C. A., Gómez, C. A., Mizuno, Y., Arnsten, J. H., Wilkinson, J. D., Knight, K. R., Knowlton, A. R., Santibanez, S., Tobin, K. E., Rose, C. D., Valverde, E. E., Gourevitch, M. N., Eldred, L., Borkowf, C. B., & INSPIRE Study Team (2007). Results from a randomized controlled trial of a peer-mentoring intervention to reduce HIV transmission and increase access to care and adherence to HIV medications among HIV-seropositive injection drug users. Journal of acquired immune deficiency syndromes (1999), 46 *Suppl 2*, S35–S47.

Pyne, J. M., Fortney, J. C., Curran, G. M., Tripathi, S., Atkinson, J. H., Kilbourne, A. M., Hagedorn, H. J., Rimland, D., Rodriguez-Barradas, M. C., Monson, T., Bottonari, K. A., Asch, S. M., & Gifford, A. L. (2011). Effectiveness of collaborative care for depression in human immunodeficiency virus clinics. Archives of internal medicine, 171(1), 23–31.

Rawlings, M. K., Thompson, M. A., Farthing, C. F., Brown, L. S., Racine, J., Scott, R. C., Crawford, K. H., Goodwin, S. D., Tolson, J. M., Williams, V. C., Shaefer, M. S., & NZTA4006 Helping to Enhance Adherence to Antiretroviral Therapy (HEART) Study Team (2003). Impact of an educational program on efficacy and adherence with a twice-daily lamivudine/zidovudine/abacavir regimen in underrepresented HIV-infected patients. Journal of acquired immune deficiency syndromes (1999), *34*(2), 174–183.

Reif, L. K., Abrams, E. J., Arpadi, S., Elul, B., McNairy, M. L., Fitzgerald, D. W., & Kuhn, L. (2020). Interventions to Improve Antiretroviral Therapy Adherence Among Adolescents and Youth in Low- and Middle-Income Countries: A Systematic Review 2015-2019. AIDS and behavior, 24(10), 2797–2810.

Remien, R. H., Stirratt, M. J., Dolezal, C., Dognin, J. S., Wagner, G. J., Carballo-Dieguez, A., El-Bassel, N., & Jung, T. M. (2005). Couple-focused support to improve HIV medication adherence: a randomized controlled trial. AIDS (London, England), 19(8), 807–814.

Rich, M. L., Miller, A. C., Niyigena, P., Franke, M. F., Niyonzima, J. B., Socci, A., Drobac, P. C., Hakizamungu, M., Mayfield, A., Ruhayisha, R., Epino, H., Stulac, S., Cancedda, C., Karamaga, A., Niyonzima, S., Yarbrough, C., Fleming, J., Amoroso, C., Mukherjee, J., Murray, M., … Binagwaho, A. (2012). Excellent clinical outcomes and high retention in care among adults in a community-based HIV treatment program in rural Rwanda. Journal of acquired immune deficiency syndromes (1999), 59(3), e35–e42.

Robbins, G. K., Testa, M. A., Su, M., Safren, S. A., Morse, G., Lammert, S., Shafer, R. W., Reynolds, N. R., & Chesney, M. A. (2013). Site nurse-initiated adherence and symptom support telephone calls for HIV-positive individuals starting antiretroviral therapy, ACTG 5031: substudy of ACTG 384. HIV clinical trials, 14(5), 235–253.

Roberts K. J. (2000). Barriers to and facilitators of HIV-positive patients’ adherence to antiretroviral treatment regimens. AIDS patient care and STDs, 14(3), 155–168.

Rosen, J. G., Nakyanjo, N., Isabirye, D., Wawer, M. J., Nalugoda, F., Reynolds, S. J., Nakigozi, G., Grabowski, M. K., & Kennedy, C. E. (2020). Antiretroviral treatment sharing among highly mobile Ugandan fisherfolk living with HIV: a qualitative study. AIDS care, 32(7), 912–915.

Rosen, M. I., Dieckhaus, K., McMahon, T. J., Valdes, B., Petry, N. M., Cramer, J., & Rounsaville, B. (2007). Improved adherence with contingency management. AIDS patient care and STDs, 21(1), 30–40.

Roy Paladhi, U., Katz, D. A., Farquhar, C., & Thirumurthy, H. (2022). Using Behavioral Economics to Support PrEP Adherence for HIV Prevention. Current HIV/AIDS reports, 19(5), 409–414.

Ruiz, I., Olry, A., López, M. A., Prada, J. L., & Causse, M. (2010). Prospective, randomized, two-arm controlled study to evaluate two interventions to improve adherence to antiretroviral therapy in Spain. Enfermedades infecciosas y microbiologia clinica, 28(7), 409–415.

Saberi, P., Dong, B. J., Johnson, M. O., Greenblatt, R. M., & Cocohoba, J. M. (2012). The impact of HIV clinical pharmacists on HIV treatment outcomes: a systematic review. Patient preference and adherence, 6, 297–322.

Sabin, L. L., DeSilva, M. B., Hamer, D. H., Xu, K., Zhang, J., Li, T., Wilson, I. B., & Gill, C. J. (2010). Using electronic drug monitor feedback to improve adherence to antiretroviral therapy among HIV-positive patients in China. AIDS and behavior, 14(3), 580–589.

Safren, S. A., O’Cleirigh, C. M., Bullis, J. R., Otto, M. W., Stein, M. D., & Pollack, M. H. (2012). Cognitive behavioral therapy for adherence and depression (CBT-AD) in HIV-infected injection drug users: a randomized controlled trial. Journal of consulting and clinical psychology, 80(3), 404–415.

Samet, J. H., Horton, N. J., Meli, S., Dukes, K., Tripps, T., Sullivan, L., & Freedberg, K. A. (2005). A randomized controlled trial to enhance antiretroviral therapy adherence in patients with a history of alcohol problems. Antiviral therapy, 10(1), 83–93.

Sarna, A., Luchters, S., Geibel, S., Chersich, M. F., Munyao, P., Kaai, S., Mandaliya, K. N., Shikely, K. S., Temmerman, M., & Rutenberg, N. (2008). Short- and long-term efficacy of modified directly observed antiretroviral treatment in Mombasa, Kenya: a randomized trial. Journal of acquired immune deficiency syndromes (1999), 48(5), 611–619.

Shand, T., Thomson-de Boor, H., van den Berg, W., Peacock, D., Pascoe, L. (2014). The HIV blind spot: men and HIV testing, treatment and care in sub-Saharan Africa. IDS Bulletin. 45(1):53–60.

Shannon, K., Leiter, K., Phaladze, N., Hlanze, Z., Tsai, A. C., Heisler, M., Iacopino, V., & Weiser, S. D. (2012). Gender inequity norms are associated with increased male-perpetrated rape and sexual risks for HIV infection in Botswana and Swaziland. PloS one, 7(1), e28739.

Sharma, M., Barnabas, R. V., & Celum, C. (2017). Community-based strategies to strengthen men’s engagement in the HIV care cascade in sub-Saharan Africa. PLoS medicine, 14(4), e1002262.

Shushtari, Z. J., Salimi, Y., Sajjadi, H., & Paykani, T. (2023). Effect of Social Support Interventions on Adherence to Antiretroviral Therapy Among People Living with HIV: A Systematic Review and Meta-Analysis. AIDS and behavior, 27(5), 1619–1635.

Sikweyiya, Y. M., Jewkes, R., & Dunkle, K. (2014). Impact of HIV on and the constructions of masculinities among HIV-positive men in South Africa: implications for secondary prevention programs. Global health action, 7, 24631.

Sileo, K. M., Wanyenze, R. K., Mukasa, B., Musoke, W., & Kiene, S. M. (2021). The Intersection of Inequitable Gender Norm Endorsement and HIV Stigma: Implications for HIV Care Engagement for Men in Ugandan Fishing Communities. AIDS and behavior, 25(9), 2863– 2874.

Sileo, K. M., Miller, A. P., Huynh, T. A., & Kiene, S. M. (2020). A systematic review of interventions for reducing heavy episodic drinking in sub-Saharan African settings. PloS one, 15(12), e0242678.

Sileo, K. M., Kizito, W., Wanyenze, R. K., Chemusto, H., Musoke, W., Mukasa, B., & Kiene, S. M. (2019a). A qualitative study on alcohol consumption and HIV treatment adherence among men living with HIV in Ugandan fishing communities. AIDS care, 31(1), 35–40.

Sileo, K. M., Kizito, W., Wanyenze, R. K., Chemusto, H., Reed, E., Stockman, J. K., Musoke, W., Mukasa, B., & Kiene, S. M. (2019b). Substance use and its effect on antiretroviral treatment adherence among male fisherfolk living with HIV/AIDS in Uganda. PloS one, 14(6), e0216892.

Sileo, K. M., Reed, E., Kizito, W., Wagman, J. A., Stockman, J. K., Wanyenze, R. K., Chemusto, H., Musoke, W., Mukasa, B., & Kiene, S. M. (2019c). Masculinity and engagement in HIV care among male fisherfolk on HIV treatment in Uganda. Culture, health & sexuality, 21(7), 774–788.

Sileo, K. M., Wanyenze, R. K., Kizito, W., Reed, E., Brodine, S. K., Chemusto, H., Musoke, W., Mukasa, B., & Kiene, S. M. (2019d). Multi-level Determinants of Clinic Attendance and Antiretroviral Treatment Adherence Among Fishermen Living with HIV/AIDS in Communities on Lake Victoria, Uganda. AIDS and behavior, 23(2), 406–417.

Sileo, K. M., Fielding-Miller, R., Dworkin, S. L., & Fleming, P. J. (2019e). A scoping review on the role of masculine norms in men’s engagement in the HIV care continuum in sub-Saharan Africa. AIDS care, 31(11), 1435–1446.

Sileo, K. M., Kintu, M., Chanes-Mora, P., & Kiene, S. M. (2016). “Such Behaviors Are Not in My Home Village, I Got Them Here”: A Qualitative Study of the Influence of Contextual Factors on Alcohol and HIV Risk Behaviors in a Fishing Community on Lake Victoria, Uganda. AIDS and behavior, 20(3), 537–547.

Simoni, J. M., Wiebe, J. S., Sauceda, J. A., Huh, D., Sanchez, G., Longoria, V., Andres Bedoya, C., & Safren, S. A. (2013). A preliminary RCT of CBT-AD for adherence and depression among HIV-positive Latinos on the U.S.-Mexico border: the Nuevo Día study. AIDS and behavior, 17(8), 2816–2829.

Simoni, J. M., Huh, D., Frick, P. A., Pearson, C. R., Andrasik, M. P., Dunbar, P. J., & Hooton, T. M. (2009). Peer support and pager messaging to promote antiretroviral modifying therapy in Seattle: a randomized controlled trial. Journal of acquired immune deficiency syndromes (1999), *52*(4), 465–473.

Simoni, J. M., Pantalone, D. W., Plummer, M. D., & Huang, B. (2007). A randomized controlled trial of a peer support intervention targeting antiretroviral medication adherence and depressive symptomatology in HIV-positive men and women. Health psychology : official journal of the Division of Health Psychology, American Psychological Association, 26(4), 488–495.

Singh, R. J., Sarna, A., Schensul, J. J., Mahapatra, B., Ha, T., & Schensul, S. L. (2020). A multilevel intervention to reduce stigma among alcohol consuming men living with HIV receiving antiretroviral therapy: findings from a randomized control trial in India. AIDS (London, England), 34 *Suppl 1*, S83–S92.

Siu, G. E., Wight, D., & Seeley, J. (2014). ’Dented’ and ’resuscitated’ masculinities: the impact of HIV diagnosis and/or enrolment on antiretroviral treatment on masculine identities in rural eastern Uganda. SAHARA J : journal of Social Aspects of HIV/AIDS Research Alliance, 11(1), 211–221.

Siu, G. E., Seeley, J., & Wight, D. (2013). Dividuality, masculine respectability and reputation: how masculinity affects men’s uptake of HIV treatment in rural eastern Uganda. Social science & medicine (1982), 89, 45–52.

Siu, G. E., Wight, D., & Seeley, J. (2012). How a masculine work ethic and economic circumstances affect uptake of HIV treatment: experiences of men from an artisanal gold mining community in rural eastern Uganda. Journal of the International AIDS Society, 15 Suppl 1(Suppl 1), 1–9.

Sorensen, J. L., Haug, N. A., Delucchi, K. L., Gruber, V., Kletter, E., Batki, S. L., Tulsky, J. P., Barnett, P., & Hall, S. (2007). Voucher reinforcement improves medication adherence in HIV-positive methadone patients: a randomized trial. Drug and alcohol dependence, 88(1), 54–63.

Ssewamala, F. M., Neilands, T. B., Waldfogel, J., & Ismayilova, L. (2012). The impact of a comprehensive microfinance intervention on depression levels of AIDS-orphaned children in Uganda. The Journal of adolescent health : official publication of the Society for Adolescent Medicine, 50(4), 346–352.

Strathdee, S. A., Shoptaw, S., Dyer, T. P., Quan, V. M., Aramrattana, A., & Substance Use Scientific Committee of the HIV Prevention Trials Network (2012). Towards combination HIV prevention for injection drug users: addressing addictophobia, apathy and inattention. Current opinion in HIV and AIDS, 7(4), 320–325.

Thiede, H., Jenkins, R. A., Carey, J. W., Hutcheson, R., Thomas, K. K., Stall, R. D., White, E., Allen, I., Mejia, R., & Golden, M. R. (2009). Determinants of recent HIV infection among Seattle-area men who have sex with men. American journal of public health, 99 Suppl 1(Suppl 1), S157–S164.

Thurman, T. R., Haas, L. J., Dushimimana, A., Lavin, B., & Mock, N. (2010). Evaluation of a case management program for HIV clients in Rwanda. AIDS care, 22(6), 759–765.

Tiberio, J., Laurent, Y. I., Ndayongeje, J., Msami, A., Welty, S., Ngonyani, A., Mwankemwa, S., Makumbuli, M., McFarland, W., & Morris, M. D. (2018). Context and characteristics of illicit drug use in coastal and interior Tanzania. The International journal on drug policy, 51, 20–26.

Tirivayi, N., & Groot, W. (2011). Health and welfare effects of integrating AIDS treatment with food assistance in resource constrained settings: a systematic review of theory and evidence. Social science & medicine (1982), 73(5), 685–692.

Tsondai, P. R., Wilkinson, L. S., Grimsrud, A., Mdlalo, P. T., Ullauri, A., & Boulle, A. (2017). High rates of retention and viral suppression in the scale-up of antiretroviral therapy adherence clubs in Cape Town, South Africa. Journal of the International AIDS Society, 20(Suppl 4), 21649.

Tuldrà, A., Fumaz, C. R., Ferrer, M. J., Bayés, R., Arnó, A., Balagué, M., Bonjoch, A., Jou, A., Negredo, E., Paredes, R., Ruiz, L., Romeu, J., Sirera, G., Tural, C., Burger, D., & Clotet, B. (2000). Prospective randomized two-Arm controlled study to determine the efficacy of a specific intervention to improve long-term adherence to highly active antiretroviral therapy. Journal of acquired immune deficiency syndromes (1999), *25*(3), 221–228.

Tumwesigye, N. M., Atuyambe, L., Wanyenze, R. K., Kibira, S. P., Li, Q., Wabwire-Mangen, F., & Wagner, G. (2012). Alcohol consumption and risky sexual behaviour in the fishing communities: evidence from two fish landing sites on Lake Victoria in Uganda. BMC public health, 12, 1069.

UNAIDS, 2022. Global HIV & AIDS statistics - Fact sheet https://www.unaids.org/en/resources/fact-sheet (accessed on 20 March 2023).

UNAIDS (2018). Fact sheet—Latest statistics on the status of the AIDS epidemic. Available online: http://www.unaids.org/en/resources/fact-sheet (accessed on 20 March 2023).

UNAIDS (2014). The Gap Report. Geneva: https://www.hivlawandpolicy.org/resources/gap-report-joint-united-nations-programme-hivaids-unaids-2014 (accessed on 30 March 2023)

Uzma, Q., Emmanuel, F., Ather, U., & Zaman, S. (2011). Efficacy of Interventions for Improving Antiretroviral Therapy Adherence in HIV/AIDS Cases at PIMS, Islamabad. Journal of the International Association of Physicians in AIDS Care (Chicago, Ill.: 2002), 10(6), 373–383.

Vella, S., Schwartländer, B., Sow, S. P., Eholie, S. P. & Murphy, R. L. (2012). The history of antiretroviral therapy and of its implementation in resource-limited areas of the world. AIDS, 26 (10), 1231–1241. doi: 10.1097/QAD.0b013e32835521a3.

Walter, K. N., & Petry, N. M. (2016). Lifetime suicide attempt history, quality of life, and objective functioning among HIV/AIDS patients with alcohol and illicit substance use disorders. International journal of STD & AIDS, 27(6), 476–485.

Wandera, S. O., Tumwesigye, N. M., Walakira, E. J., Kisaakye, P., & Wagman, J. (2021). Alcohol use, intimate partner violence, and HIV sexual risk behavior among young people in fishing communities of Lake Victoria, Uganda. BMC public health, 21(1), 544.

Wang, H., Zhou, J., Huang, L., Li, X., Fennie, K. P., & Williams, A. B. (2010). Effects of nurse-delivered home visits combined with telephone calls on medication adherence and quality of life in HIV-infected heroin users in Hunan of China. Journal of clinical nursing, 19(3-4), 380–388.

Weber, R., Ruppik, M., Rickenbach, M., Spoerri, A., Furrer, H., Battegay, M., Cavassini, M., Calmy, A., Bernasconi, E., Schmid, P., Flepp, M., Kowalska, J., Ledergerber, B., & Swiss HIV Cohort Study (SHCS) (2013). Decreasing mortality and changing patterns of causes of death in the Swiss HIV Cohort Study. HIV medicine, 14(4), 195–207.

Weber, R., Christen, L., Christen, S., Tschopp, S., Znoj, H., Schneider, C., Schmitt, J., Opravil, M., Günthard, H. F., Ledergerber, B., & Swiss HIV Cohort Study (2004). Effect of individual cognitive behaviour intervention on adherence to antiretroviral therapy: prospective randomized trial. Antiviral therapy, 9(1), 85–95.

West, B. S., Choo, M., El-Bassel, N., Gilbert, L., Wu, E., & Kamarulzaman, A. (2014). Safe havens and rough waters: networks, place, and the navigation of risk among injection drug-using Malaysian fishermen. The International journal on drug policy, 25(3), 575–582.

WHO, (November 2022). HIV Key Facts. https://www.who.int/news-room/fact-sheets/detail/hiv-aids#:~:text=HIV%20continues%20to%20be%20a,no%20cure%20for%20HIV%infection (accessed on 22 March 2023).

WHO (2007). Engaging Men and Boys in Changing Gender-Based Inequity in Health: Evidence from Programme Interventions. Geneva: World Health Organization. https://apps.who.int/iris/bitstream/handle/10665/43679/9789241595490_eng.pdf (accessed on 1 April 2023).

WHO (2003). Adherence to long-term therapies: Evidence for action. http://www.who.int/chp/knowledge/publications/adherence_report/en/ (accessed on 2 May 2023).

Witte, S. S., Aira, T., Tsai, L. C., Riedel, M., Offringa, R., Chang, M., El-Bassel, N., & Ssewamala, F. (2015). Efficacy of a savings-led microfinance intervention to reduce sexual risk for HIV among women engaged in sex work: a randomized clinical trial. American journal of public health, 105(3), e95–e102.

Womack, J. A., & Justice, A. C. (2020). The OATH Syndemic: opioids and other substances, aging, alcohol, tobacco, and HIV. Current opinion in HIV and AIDS, 15(4), 218–225.

Woolf-King, S. E., & Maisto, S. A. (2011). Alcohol use and high-risk sexual behavior in Sub-Saharan Africa: a narrative review. Archives of sexual behavior, 40(1), 17–42.

