## Supplemental files for "Antiretroviral therapy adherence interventions among persons who use alcohol and other substances in fisherfolk communities: A systematic review"

**Supplementary Files - Appendices**

**Appendix 1: Sample of search strategy and electronic database pilot**

PubMed

("substance related disorders"[MeSH Terms] OR "drug misuse"[MeSH Terms]) AND ("fisheries"[MeSH Terms] OR "fisherfolk communit*"[Text Word] OR "fishing communit*"[Text Word] OR "fishing zone"[Text Word] OR "fishing village"[Text Word] OR "fish farm"[Text Word] OR "fishing area"[Text Word] OR "fisherfolk"[Text Word] OR "fishermen"[Text Word] OR "fisherwomen"[Text Word]) AND ("behavior therapy"[MeSH Terms] OR "psychosocial intervention"[MeSH Terms] OR "treatment"[Text Word] OR "therapeutics"[MeSH Terms] OR "patient care"[MeSH Terms] OR "encourage"[Text Word] OR "adherence"[Text Word] OR "promote"[Text Word] OR "treatment adherence and compliance"[MeSH Terms] OR "medication adherence"[MeSH Terms] OR "anti retroviral therapy"[Text Word] OR "hiv infections"[MeSH Terms] OR "acquired immunodeficiency syndrome"[MeSH Terms] OR "antiretroviral therapy, highly active"[MeSH Terms] OR "prevent hiv transmission"[Text Word] OR "health education"[MeSH Terms] OR "health promotion"[MeSH Terms] OR "harm reduction"[MeSH Terms])

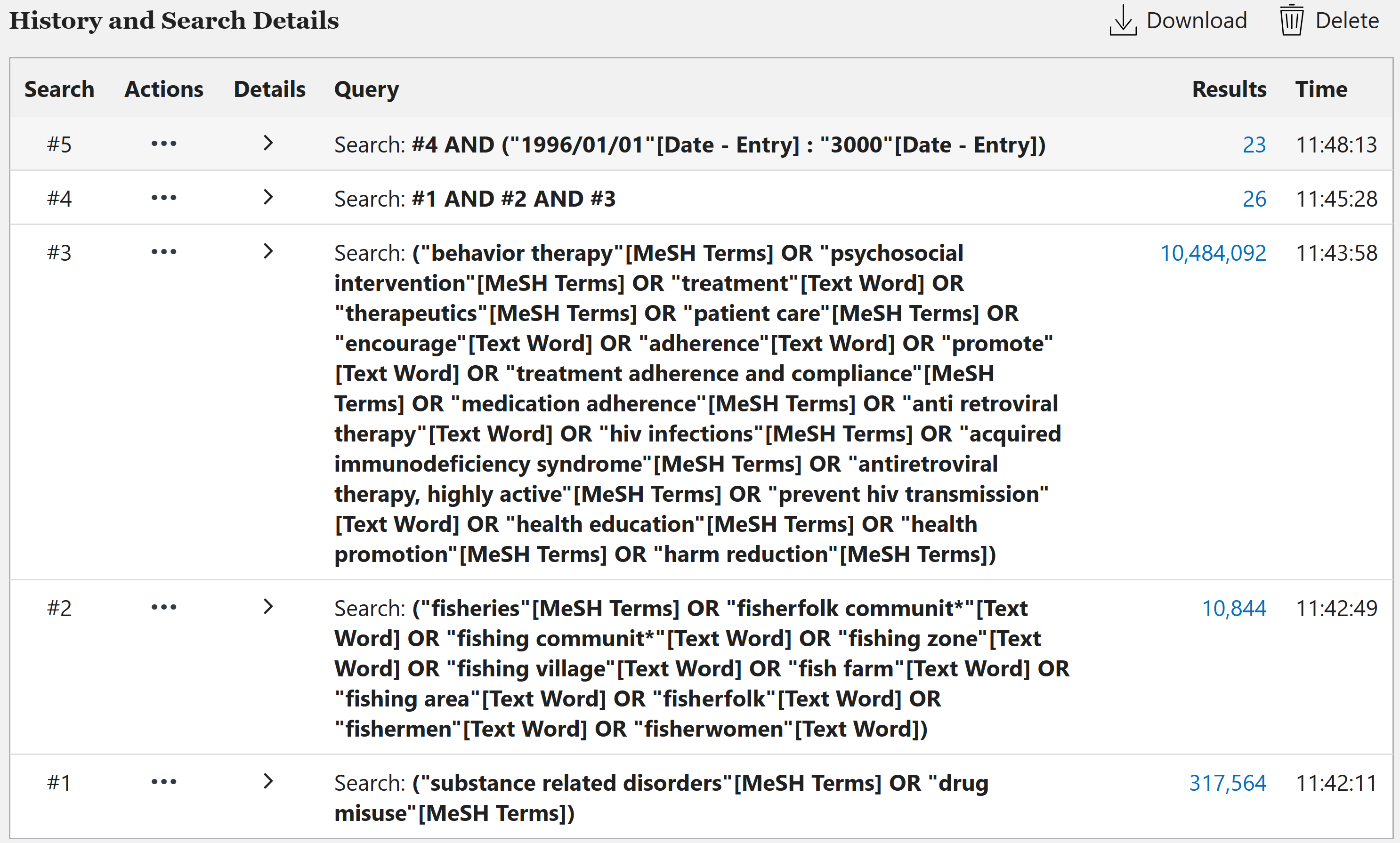

**Appendix 2: Characteristics of excluded studies**

| **Study 1D** | **Rationale for exclusion** |
| --- | --- |
| Bonnevie et al., 2020 | Respondents include age below 18 years and outcomes are not segregated by age |
| Burgos-Soto et al., 2020 | Respondents include age below 18 years and outcomes are not segregated by age |
| Kuteesa et al., (2022) | Respondents include age below 18 years and outcomes are not segregated by age |
| Lubega et al., 2015 | Respondents include age below 18 years and outcomes are not segregated by age |
| Mgabo et al., 2013 | Respondents include age below 18 years and outcomes are not segregated by age |
| Burke et al., 2017 | No alcohol or other substance use |
| Chang et al., 2016 | No alcohol or other substance use |
| Kagaayi et al., 2019 | No alcohol or other substance use |
| Long et al., 2017 | No alcohol or other substance use |
| Rosen et al., 2020 | No alcohol or other substance use |
| Sileo et al., (2019d) | No alcohol or other substance use |
| Brown et al., 2017 | No ART adherence intervention outcome or to encourage adherence |
| Kiene et al., 2019b | No ART adherence intervention outcome or to encourage adherence |
| Kissling et al., 2005 | No ART adherence intervention outcome or to encourage adherence |
| Ousley et al., 2018 | No ART adherence intervention outcome or to encourage adherence |
| Brown & George, 2019 | No ART adherence intervention outcome or to encourage adherence |
| Hickey et al., 2015 | Study not conducted in fisherfolk community |
